# *BIRC6* modifies risk of invasive bacterial infection in Kenyan children

**DOI:** 10.1101/2022.02.18.22271173

**Authors:** James J Gilchrist, Silvia Kariuki, James A Watson, Gavin Band, Sophie Uyoga, Carolyne M Ndila, Neema Mturi, Salim Mwarumba, Shebe Mohammed, Moses Mosobo, Kirk A Rockett, Alexander J Mentzer, Dominic P Kwiatkowski, Adrian VS Hill, Kathryn Maitland, J Anthony G Scott, Thomas N Williams

## Abstract

Invasive bacterial disease is a major cause of morbidity and mortality in African children. Despite being caused by diverse pathogens, children with sepsis are clinically indistinguishable from one another. In spite of this, most genetic susceptibility loci for invasive infection that have been discovered to date are pathogen specific and are not therefore suggestive of a shared genetic architecture of bacterial sepsis. Here we utilise probabilistic diagnostic models to identify children with a high probability of invasive bacterial disease among critically unwell Kenyan children with *P. falciparum* parasitaemia. We construct a joint dataset including 1,445 bacteraemia cases and 1,143 severe malaria cases, and population controls, among critically unwell Kenyan children that have previously been genotyped for human genetic variation. Using these data we perform a cross-trait genome-wide association study of invasive bacterial infection, weighting cases according to their probability of bacterial disease. In doing so we identify and validate a novel risk locus for invasive infection secondary to multiple bacterial pathogens, that has no apparent effect on malaria risk. The locus identified modifies splicing of *BIRC6* in stimulated monocytes, implicating regulation of apoptosis and autophagy in the pathogenesis of sepsis in Kenyan children.

## Introduction

Invasive bacterial diseases are a major cause of child morbidity and mortality in Africa (***Berkley et al., 2005***). Although improved control measures, including the rollout of anti-bacterial conjugate vaccines (***Cowgill et al., 2006***; ***Silaba et al., 2019***), have led to recent declines, the burden of conditions such as pneumonia, meningitis and sepsis secondary to bacterial pathogens remains significant (***Vos et al., 2020***). A better understanding of the biology of invasive bacterial infections in African populations might help the development of novel interventions.

Susceptibility to invasive bacterial infections varies widely between individuals. In African children, some of this variability is determined by acquired comorbidities such as HIV, malnutrition and malaria (***Berkley et al., 2005***; ***Church and Maitland, 2014***; ***Scott et al., 2011***). However, the identification of common genetic variants as determinants of bacterial infection suggests that a significant portion of this variability is heritable. Many of these genetic susceptibility loci have pathogen-specific effects (***Davila et al., 2010***; ***Gilchrist et al., 2018***; ***Rautanen et al., 2016***), which is consistent with our understanding of infection susceptibility derived from primary immunodeficiencies. Key examples of pathogen-specificity among primary immunodeficiencies include Mendelian susceptibility to mycobacterial disease and susceptibility to non-tuberculous mycobacteria and nontyphoidal *Salmonella* (***Bustamante et al., 2014***), terminal complement deficiencies and meningococcal disease (***Figueroa et al., 1993***), and IRAK4 deficiency and pneumococcal disease (***Picard et al., 2007***). A major exception to this is the rs334 A>T mutation in *HBB* (sickle haemoglobin), which is associated with raised and lowered risks of infection secondary to a broad range of pathogens among homozygotes (***Williams et al., 2009***) and heterozygotes (***Scott et al., 2011***) respectively. However, the effect sizes associated with sickle haemoglobin are extreme, being maintained in populations by balancing selection, and larger sample sizes will probably be needed for the discovery of new variants with multi-pathogen effects.

Because the clinical features of invasive bacterial infections and severe malaria are broadly similar (***Bejon et al., 2007***), it can be difficult to distinguish between severe illness caused by extensive sequestration of malaria parasites in the microvasculature (‘true’ severe malaria) and bacterial sepsis in the presence of incidental parasitaemia on the basis of clinical features alone. This is made harder still by the fact that antibiotic pre-treatment and inadequate blood-culture volumes mean that, even in settings with excellent diagnostic facilities, true invasive bacterial infections can often not be confirmed (***Driscoll et al., 2017***). Recently, we illustrated this clinical complexity through a study in which we used probabilistic models based on malaria-specific biomarkers to show that approximately one third of children recruited to studies in Africa with a clinical diagnosis of severe malaria were actually suffering from other conditions (***Watson et al., 2021a***,b).

In the current study, we extend this work to show that invasive bacterial infections are common in children admitted to hospital with a clinical diagnosis of severe malaria, but in whom biomarkers subsequently suggest that malaria was probably not the primary cause for their severe illness. We then construct a dataset of genome-wide genotyped samples from 5,400 Kenyan children, comprising critically unwell Kenyan children with bacteraemia (***Rautanen et al., 2016***) and severe malaria (***Band et al., 2019***), and population controls. Using this dataset we perform a GWAS of invasive bacterial infection in Kenyan children, weighting cases according to the probability that their disease was mediated by a bacterial pathogen. In doing so, we increase our study power and identify *BIRC6* as a novel genetic determinant of invasive bacterial disease in Kenyan children.

## Results

### Severe malaria probability and risk of bacteraemia

Children admitted to the high dependency ward of Kilifi County Hospital with a clinical diagnosis of severe malaria, defined as a severe febrile illness in the presence of *P. falciparum* parasitaemia (n=2,200), between 11th June 1995 and 12th June 2008 were included in the study. While this definition is sensitive it is not specific, meaning that our study will have included some children with sepsis accompanied by incidental parasitaemia (***Watson et al., 2021a***). We therefore used two probabilistic models, which included either platelet counts and plasma *Pf* HRP2 concentrations (Model 1, n=1,400) or white blood cell and platelet counts (Model 2, n=2,200), to determine the likelihood of ‘true’ severe malaria among these children. The estimated probabilities of ‘true’ severe malaria using each model were well-correlated (*r* = 0.64). Of 1,400 children with a clinical diagnosis of severe malaria with measured plasma *Pf* HRP2 concentrations, 425 (30.4%, Figure 1A and 1B) had a low probability (P(SM|Data)<0.5) of having ‘true’ severe malaria (941 of 2,220 children using WBC and platelet count data, Figure 1–figure supplement 1A and 1B). That is, while they presented with febrile illness and concomitant malaria parasitaemia, it is unlikely that their illnesses were directly attributable to malaria.

**Figure 1.**
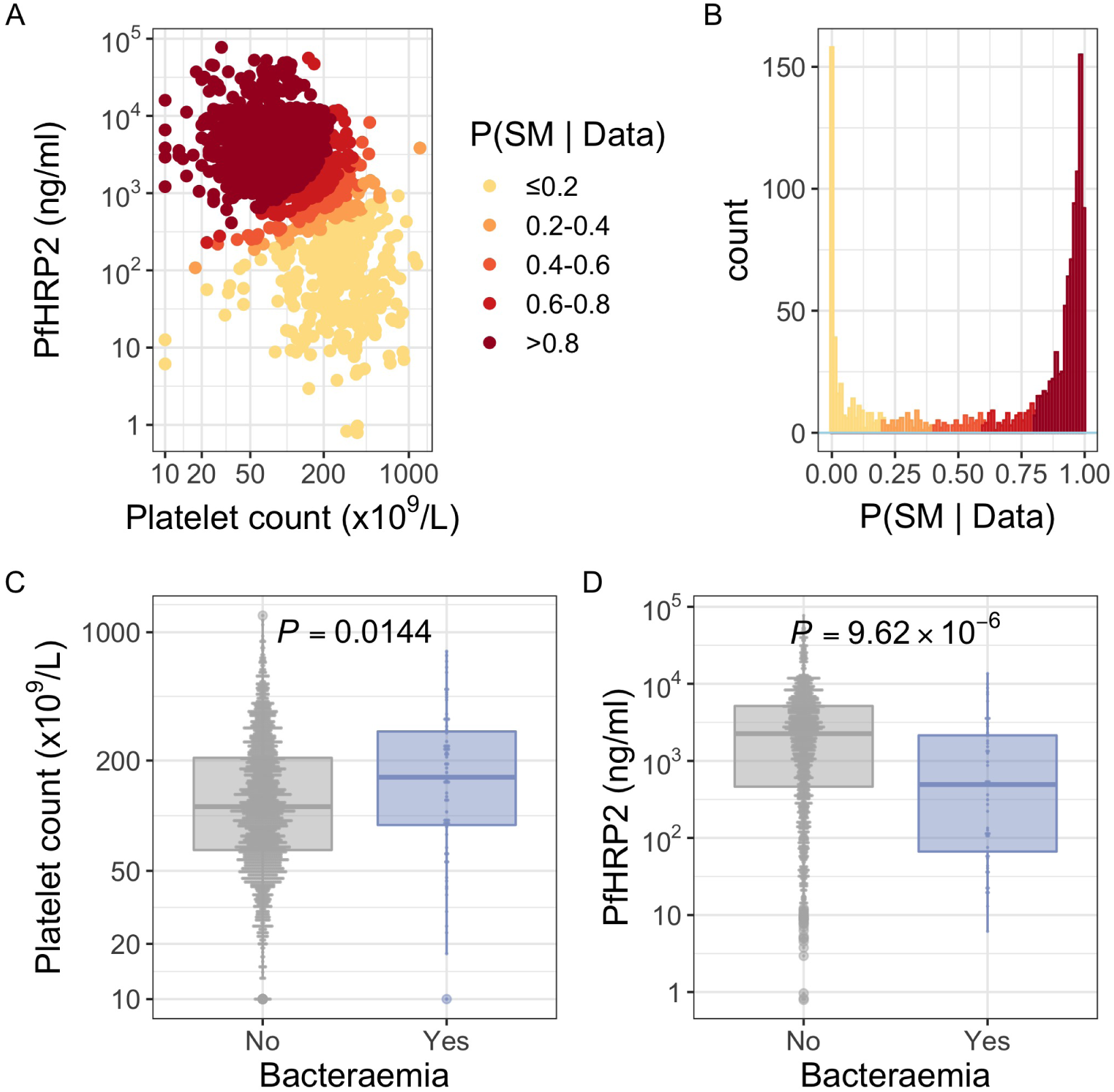
*Pf* HRP2 and platelet count as predictors of severe malaria. (A) Distribution of *Pf* HRP2 concentrations and platelet count among Kenyan children (n=1,400) with a clinical diagnosis of severe malaria. Points are coloured according to the probability of ‘true’ severe malaria given the data. (B) Distribution of ‘true’ severe malaria probabilities estimated from platelet count and plasma *Pf* HRP2 concentrations. (C) Platelets counts in children with a clinical diagnosis of severe malaria with and without concomitant bacteraemia. (D) *Pf* HRP2 concentrations in children with a clinical diagnosis of severe malaria with and without concomitant bacteraemia.

In keeping with the hypothesis that a significant proportion of these critically unwell children represented culture-negative invasive bacterial disease (Figure 1), in-patient mortality was higher among children with a low than a high probability of ‘true’ severe malaria (Table 1; *OR*_*model*1_ = 1.57, 95% CI 1.11 − 2.21, *p* = 0.01; *OR*_*model*2_ = 2.09, 95% CI 1.60 − 2.72, *p* = 4.91 × 10^−8^). This was also reflected in the rates of concurrent bacteraemia (Table 1; *OR*_*model*1_ = 2.92, 95% CI 1.66 − 5.13, *p* = 1.07 × 10^−4^; *OR*_*model*2_ = 2.00, 95% CI 1.27 − 3.17, *p* = 0.003). Similarly, the constituents of model 1 were each associated with blood culture positivity, both higher platelet counts (OR=2.36, 95% CI 1.19-4.70, *p* = 0.014) and lower *Pf* HRP2 levels (OR=0.52, 95% CI 0.39-0.70, *p* = 9.62 × 10^−^6) both being associated with the risk of coincident bacteraemia (Figure 1C and 1D). Conversely, white blood counts in isolation were not associated with risk of concurrent bacteraemia (Figure 1–figure supplement 1). Plasma *Pf* HRP2 is the single best biomarker for severe malaria (***Hendriksen et al., 2012***). In light of this, and given the greater enrichment for concurrent bacteraemia among children with a low probability of ‘true’ severe malaria as calculated by Model 1 than Model 2, we used Model 1 probabilities in downstream analyses where available (n=1,400) and used Model 2 probabilities for all other cases (n=800).

**Table 1.** Demographics & clinical characteristics of Kenyan children with severe malaria.

| Model | Numbers | Sex (female) | Age (months) | Bacteraemia | Mortality |
| --- | --- | --- | --- | --- | --- |
| *Pf*HRP2/Plt | Total (n=1,400) | 695 (49.6%) | 29 (17-44) | 51 (3.6%) | 155 (11.1%) |
|  | P(SM\|Data)>0.5 (n=975) | 497 (51.0%) | 29 (18-45) | 23 (2.4%) | 94 (7.4%) |
|  | P(SM\|Data)<0.5 (n=425) | 198 (46.6%) | 28 (16-43) | 28 (6.6%) | 61 (14.4%) |
| WBC/Plt | Total (n=2,220) | 1,074 (48.4%) | 28 (15-43) | 78 (3.5%) | 256 (11.6%) |
|  | P(SM\|Data)>0.5 (n=1,279) | 623 (48.7%) | 29 (17-44) | 32 (2.5%) | 106 (8.4%) |
|  | P(SM\|Data)<0.5 (n=941) | 451 (47.9%) | 25 (13-40) | 46 (4.9%) | 150 (15.9%) |

Mortality reflects in-patient deaths. Figures are absolute numbers with percentages or interquartile ranges in parentheses. P(SM|Data) reflects the probability of 'true' severe malaria estimated from each model (*Pf*HRP2/Platelet count, White blood cell count/Platelet count).

### GWAS of invasive bacterial disease in Kenyan children

Children with a clinical diagnosis of severe malaria but a low probability of having ‘true’ severe malaria, are thus enriched for invasive bacterial disease. Motivated by this observation we performed a genome-wide association study of invasive bacterial infection in Kenyan children in which we included both children with culture-confirmed bacteraemia and children with a clinical diagnosis of severe malaria. Children admitted to Kilifi County Hospital between 1st August 1998 and 30th October 2010 with community-acquired bacteraemia were recruited to the study as well as children from the severe malaria study described above. Control children were recruited from the same population between 1st August 2006 and 30th December 2010 as described in detail previously (***Scott et al., 2011***).

Following quality control measures (see Materials and Methods), we included 1,445 cases of culture-confirmed bacteraemia, 1,143 cases of severe malaria and 2,812 control children in our current analysis (Table 2). To account for the varying proportion of invasive bacterial disease among severe malaria cases, we applied weights to our regression analysis to reflect the greater likelihood of invasive bacterial disease among children with a low probability of ‘true’ severe malaria (sample weight, *w* = 1 − *P* (*SM Data*)). Where *Pf* HRP2 concentrations were available (n=909) we used *Pf* HRP2 and platelet count to determine *P* (*SM Data*) while we used white cell and platelet counts (n=234) in cases where they were not available. Cases with culture-proven bacteraemia and control samples were assigned a sample weight of *w* = 1. Inclusion of the 6 major principal components of genotyping data and genotyping platform as covariates in the model adequately controlled for confounding variation (*λ* = 1.0208, Figure 2–figure supplement 1). In that analysis we found evidence supporting an association between risk of invasive bacterial disease in Kenyan children and 7 SNPs at a single locus on chromosome 2 (peak SNP: rs183868412:T, OR=2.13, 95% CI 1.65-2.74, *p* = 4.64 × 10^−9^) (Figure 2, Table 3). Fine-mapping of this association identified a credible set of 7 SNPs with a 95% probability of containing the causal variant (Table 3), spanning a 212kb region: chr2:32,402,640-32,614,746.

**Table 2.**
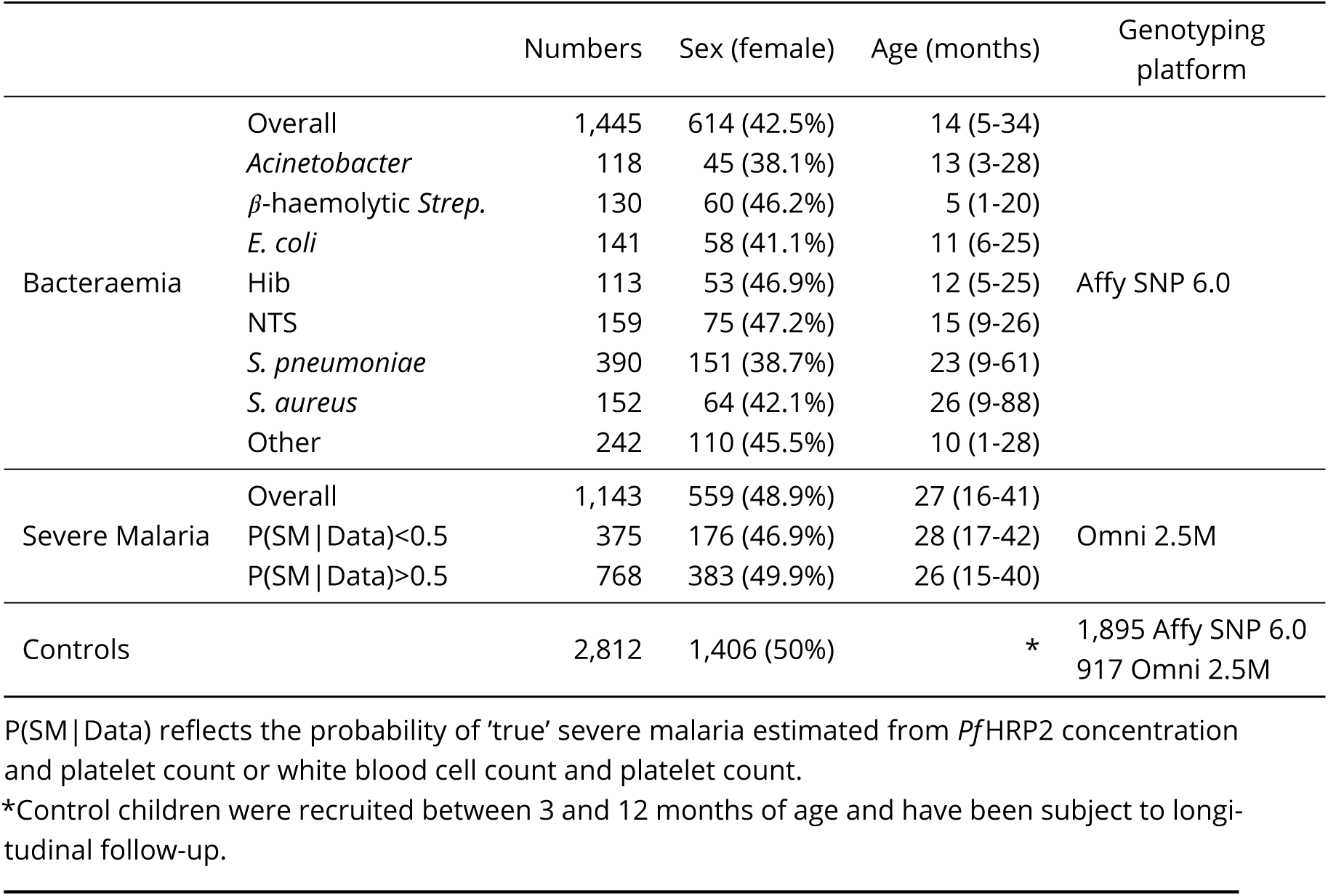
Demographics & clinical characteristics of GWAS study samples.

**Figure 2.**
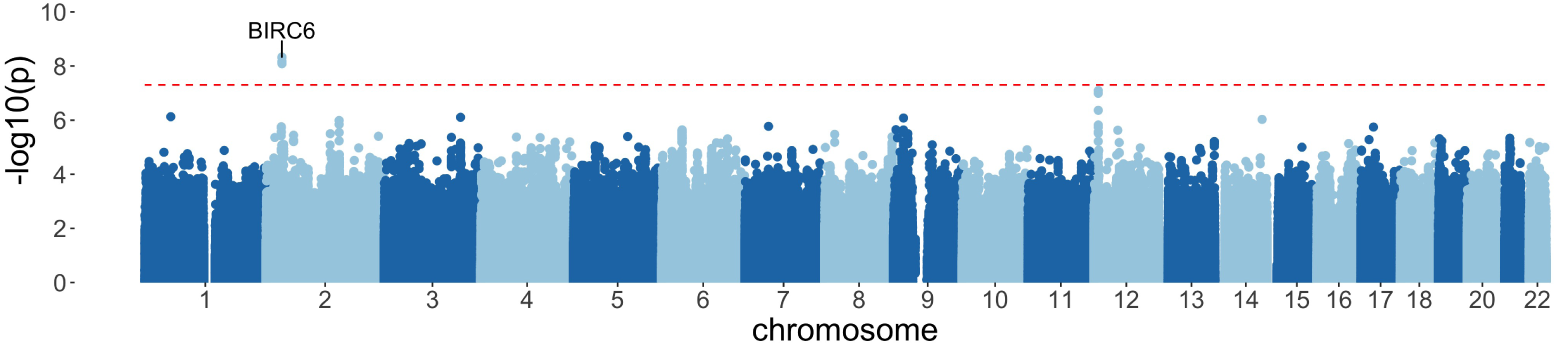
Manhattan plot of invasive bacterial infection in Kenyan children. Evidence for association with invasive bacterial disease at genotyped and imputed autosomal SNPs and indels (n=14,010,600) in Kenyan children (bacteraemia cases = 1,445, severe malaria cases = 1,143, controls = 2,812). Association statistics represent additive association. The red, dashed line denotes *p* = 5 × 10^−8^.

**Table 3.** 95% credible SNP set of invasive bacterial disease association.

| SNP | Effect allele | Chr | BP | MAF | Info score | OR (95% CI) | p-value |
| --- | --- | --- | --- | --- | --- | --- | --- |
| rs183868412 | T | 2 | 32,478,169 | 0.021 | 0.956 | 2.13 (1.65-2.74) | $4.64 \times 10^{-9}$ |
| rs139827594 | G | 2 | 32,402,640 | 0.020 | 0.966 | 2.12 (1.65-2.73) | $4.96 \times 10^{-9}$ |
| rs144257579 | G | 2 | 32,507,619 | 0.021 | 0.954 | 2.11 (1.64-2.72) | $6.82 \times 10^{-9}$ |
| rs145056232 | C | 2 | 32,503,024 | 0.021 | 0.955 | 2.11 (1.64-2.72) | $6.86 \times 10^{-9}$ |
| rs145315025 | G | 2 | 32,502,654 | 0.021 | 0.955 | 2.11 (1.64-2.72) | $6.87 \times 10^{-9}$ |
| rs143909151 | T | 2 | 32,531,452 | 0.021 | 0.962 | 2.11 (1.64-2.71) | $8.01 \times 10^{-9}$ |
| rs150430979 | T | 2 | 32,614,746 | 0.021 | 0.955 | 2.11 (1.64-2.72) | $8.18 \times 10^{-9}$ |

MAF, minor allele frequency. CI, confidence interval. Genomic coordinates are GRCh38.

We sought to replicate evidence of association in our discovery analysis through use of an independent case-control collection of Kenyan children with bacteraemia (n=434) and healthy controls (n=1,258) conducted in the same population. The peak trait-associated variants in the discovery analysis were well-imputed in the replication data (rs183868412 imputation info score = 0.84). In that analysis, we found evidence supporting the association at chromosome 2 with invasive bacterial disease (Figure 3, Table 4: rs183868412:T, OR=2.77, 95% CI 1.49-5.12, *p* = 1.29 × 10^−3^). In a fixed effects meta-analysis, rs183868412:T was strongly associated with risk of invasive bacterial disease in Kenyan children: OR=2.21, 95% CI 1.75-2.79, *p* = 2.42 × 10^−11^. That association was driven by children with culture-confirmed bacteraemia and critically unwell children with malaria parasites, but a low probability of ‘true’ severe malaria. In a stratified analysis (Figure 3, Table 4), rs183868412 was associated with culture-confirmed bacteraemia (OR=2.12, 95% CI 1.60-2.82, *p* = 1.94 ×10^−7^) and critical illness with parasitaemia and with a low probability of ‘true’ severe malaria (P(SM|Data)<0.5: OR=2.37, 95% CI 1.27-4.43, *p* = 6.82 × 10^−3^), but was not associated with risk of critical illness with a high probability of ‘true’ severe malaria (P(SM|Data)>0.5: *p* = 0.823).

**Figure 3.**
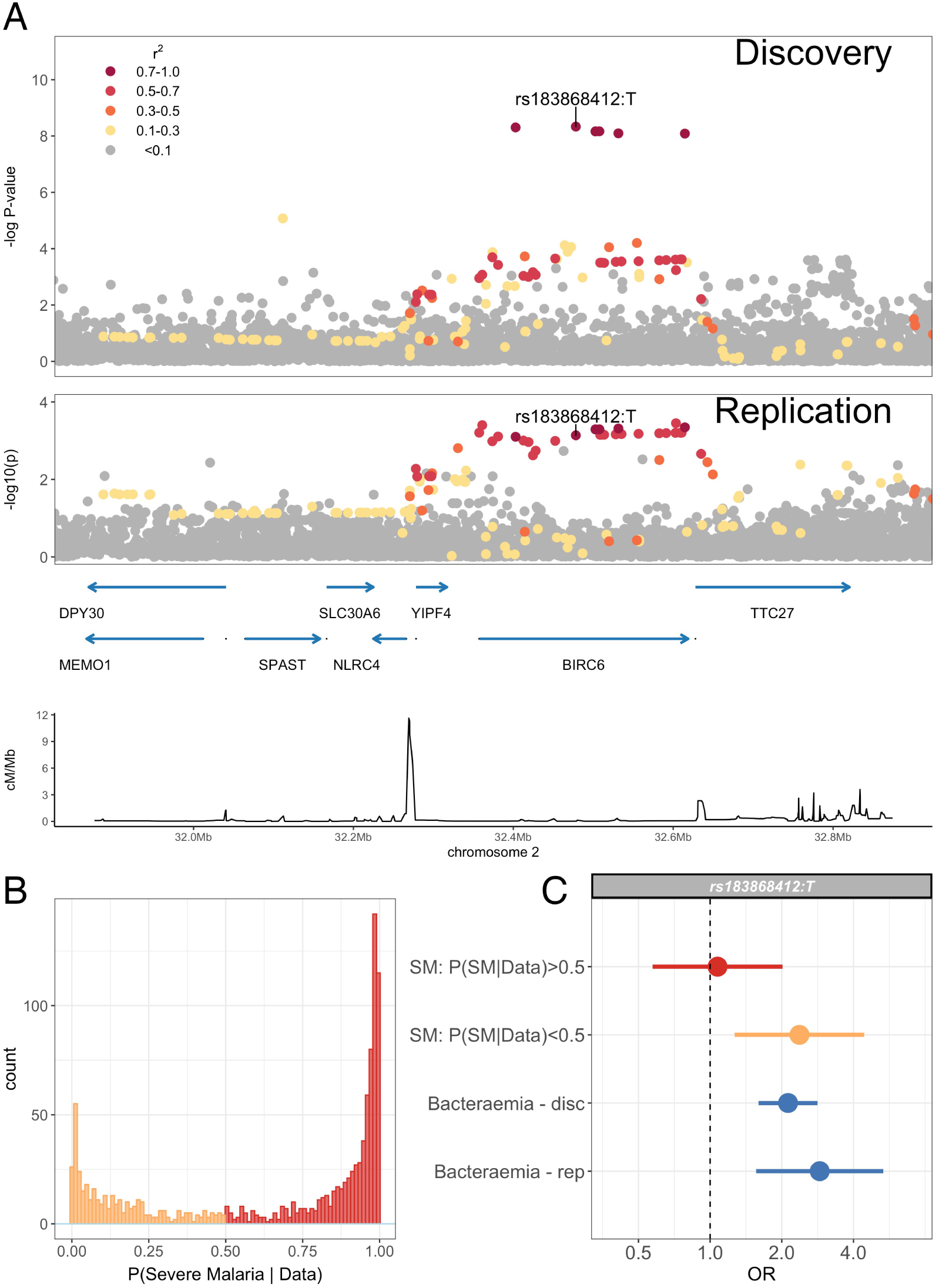
Association with invasive bacterial disease at the *BIRC6* locus. (A) Regional association plot of invasive bacterial disease association at the *BIRC6* locus in the discovery and replication analyses. SNPs are coloured according to linkage disequilibrium to rs183868412. Genomic coordinates are GRCh38. (B) Distribution of ‘true’ severe malaria probabilities among malaria cases estimated from plasma *Pf* HRP2 concentration and platelet count (n=909) and white blood cell count and platelet count (n=234). (C) Odds ratios and 95% confidence intervals of rs183868412 association with disease stratified by malaria cases with high (P>0.5, red) and low (P<0.5, orange) probabilities of ‘true’ severe malaria and culture-proven invasive bacterial disease (blue). P(SM|Data) represents the probability of ‘true’ severe malaria estimated from plasma PfHRP2 concentration and platelet count (n=909) or white blood cell count and platelet count (n=234).

**Table 4.** Effect of rs183868412 genotype on risk of invasive bacterial disease in Kenyan children.

|  |  |  | Numbers | Genotypes | MAF | OR (95% CI) | p-value |
| --- | --- | --- | --- | --- | --- | --- | --- |
| Discovery | Cases | Overall | 2,588 | 3/164/2,421 | 0.033 | 2.13 (1.65-2.74) | $p = 4.64 \times 10^{-9}$ |
| | | Bacteraemia* | 1,445 | 3/125/1,317 | 0.045 | 2.12 (1.60-2.82) | $p = 1.94 \times 10^{-7}$ |
| | | SM - P(SM\|Data)<0.5* | 375 | 0/20/355 | 0.027 | 2.37 (1.27-4.43) | $p = 6.82 \times 10^{-3}$ |
| | | SM - P(SM\|Data)>0.5* | 768 | 0/19/749 | 0.012 | 1.07 (0.57-2.01) | $p = 0.823$ |
|  | Controls |  | 2,812 | 0/111/2,701 | 0.020 |  |  |
| Replication | Cases | | 434 | 0/24/410 | 0.028 | 2.77 (1.49-5.12) | $p = 1.29 \times 10^{-3}$ |
|  | Controls |  | 1,258 | 0/28/1,230 | 0.011 |  |  |
| Meta-analysis | Cases | | 3,022 | 3/188/2,831 | 0.032 | 2.21 (1.75-2.79) | $p = 2.42 \times 10^{-11}$ |
|  | Controls |  | 4,070 | 0/139/3,931 | 0.017 |  |  |

\*estimates derived from multinomial logistic regression model. P(SM|Data) represent the probability of 'true' severe malaria estimated from plasma *Pf*HRP2 concentration and platelet count (n=909) or white blood cell count and platelet count (n=234). SM, severe malaria. MAF, minor allele frequency. CI, confidence interval.

The genetic variants associated with invasive bacterial disease in Kenyan children are monomorphic in non-African populations (https://gnomad.broadinstitute.org). Within Africa, rs183868412:T is present in all 9 African populations included in the MalariaGEN consortium project (***Band et al., 2019***) (Table 5), minor allele frequencies ranging from 0.011 in The Gambia to 0.034 in Malawi. There is no evidence for within-Africa differentiation at rs183868412 (*p* = 0.601).

**Table 5.** rs183868412 frequencies in Africa.

| Population | Number | MAF |
| --- | --- | --- |
| Gambia | 2,605 | 0.011 |
| Mali | 183 | 0.021 |
| Burkina Faso | 596 | 0.009 |
| Ghana | 320 | 0.014 |
| Nigeria | 22 | 0.024 |
| Cameroon | 685 | 0.031 |
| Malawi | 1,317 | 0.034 |
| Tanzania | 402 | 0.028 |
| Kenya | 1,681 | 0.017 |

Numbers reflect healthy control samples. MAF, minor allele frequency.

### rs183868412 is associated with risk of invasive bacterial disease secondary to diverse pathogens and is independent of malaria

Previous data describing the genetic risk of invasive bacterial disease in this population have identified pathogen-specific effects. To better-understand the range of pathogens to which genetic variation at *BIRC6* modifies risk we estimated the effect of rs183868412 on the risk of bacteraemia caused by the seven most common causative pathogens within this population (Figure 4A). In that analysis, the data best-supported a model in which genotype increases risk of bacteraemia caused by a broad range of pathogens, including bacteraemia secondary to pneumococcus, nontyphoidal *Salmonellae, E. coli, β*-haemolytic *Streptococci, S. aureus* and other less common pathogens grouped as a single stratum (log10 Bayes factor= 4.72). Genotype at rs183868412 similarly modified risk of was associated with culture-confirmed bacteraemia (OR=2.12, 95% CI 1.60-2.82, *p* = 1.94 ×10^−7^) and critical illness with parasitaemia and with a low probability of ‘true’ severe malaria (P(SM|Data)<0.5: OR=2.37, 95% CI 1.27-4.43, *p* = 6.82 × 10^−3^), but was not associated with risk of critical illness with a high probability of ‘true’ severe malaria (P(SM|Data)>0.5: *p* = 0.823).

**Figure 4.**
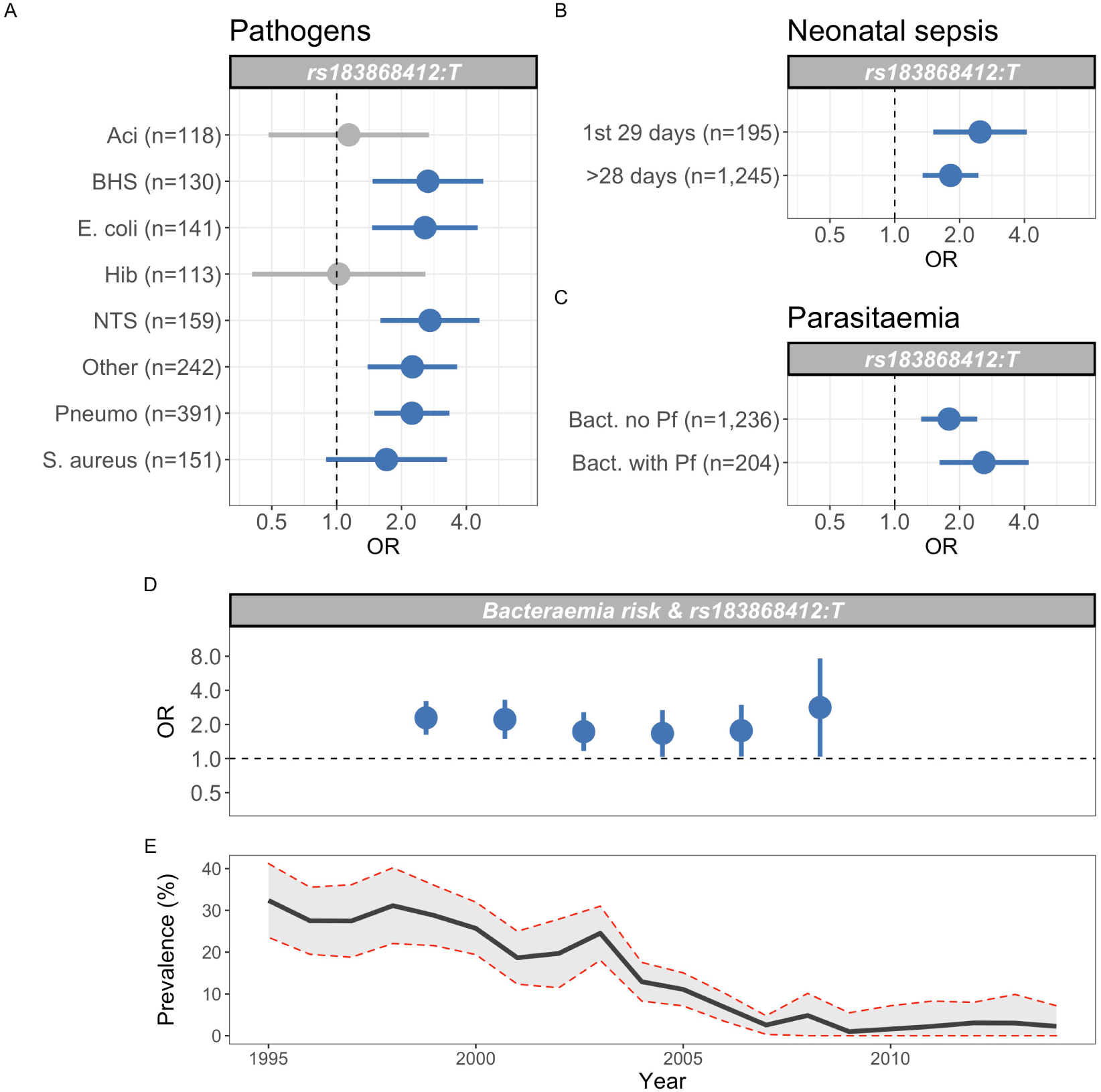
Genetic variation at *BIRC6* confers broad susceptibility to invasive bacterial disease. Odds ratios and 95% confidence intervals of rs183868412 association with invasive bacterial disease stratified by pathogen (A), neonatal and non-neonatal sepsis (B) and bacteraemia with and without malaria parasitaemia (C). Odds ratios and 95% confidence intervals of rs183868412 association with invasive bacterial disease stratified by year (D), compared to age-standardized, annual malaria parasite prevalence in Kilifi, Kenya, as estimated from parasite prevalence among trauma admissions (E). We compared models of association across strata using a Bayesian approach (see Methods). Strata associated with rs183868412 genotype in the most likely model in each analysis are highlighted in blue.

The genetic variants associated with invasive bacterial disease in Kenyan children are monomorphic in non-African populations (https://gnomad.broadinstitute.org). Within Africa, rs183868412:T is present in all 9 African populations included in the MalariaGEN consortium project (***Band et al., 2019***) (Table 5), minor allele frequencies ranging from 0.011 in The Gambia to 0.034 in Malawi. There is no evidence for within-Africa differentiation at rs183868412 (*p* = 0.601).

### rs183868412 is associated with risk of invasive bacterial disease secondary to diverse pathogens and is independent of malaria

Previous data describing the genetic risk of invasive bacterial disease in this population have identified pathogen-specific effects. To better-understand the range of pathogens to which genetic variation at *BIRC6* modifies risk we estimated the effect of rs183868412 on the risk of bacteraemia caused by the seven most common causative pathogens within this population (Figure 4A). In that analysis, the data best-supported a model in which genotype increases risk of bacteraemia caused by a broad range of pathogens, including bacteraemia secondary to pneumococcus, nontyphoidal *Salmonellae, E. coli, p*-haemolytic *Streptococci, S. aureus* and other less common pathogens grouped as a single stratum (log10 Bayes factor= 4.72). Genotype at rs183868412 similarly modified risk of bacteraemia in the neonatal period and in older children (log10 Bayes factor= 2.70, Figure 4B).

Malaria infection results in increased risk of invasive bacterial disease secondary to a broad range of pathogens (***Scott et al., 2011***), and genetic determinants of malaria risk (e.g. sickle cell trait) modify risk of bacterial infection in malaria-endemic settings (***Scott et al., 2011***). The observation that, among children with a clinical diagnosis of severe malaria, risk of disease is only modified by rs183868412 in children with a low probability that their disease represents ‘true’ severe malaria (Figure 3) suggests that the effect of genetic variation at *BIRC6* on invasive bacterial disease risk operates independently of malaria. In keeping with this, the data best-supports an effect of rs183868412 genotype on bacteraemia risk in children both with and without concomitant parasitaemia (log10 Bayes factor= 2.73, Figure 4C). In addition, unlike sickle cell trait (***Scott et al., 2011***), the increased risk of invasive bacterial infection conferred by rs183868412:T carriage in the study setting is stable across a period of declining malaria prevalence (Bayes factor= 8.7, Figure 4D).

### rs183868412 is associated with alternative splicing of *BIRC6* in stimulated monocytes

Trait associated genetic variation identified by GWAS is highly enriched for regulatory variation. The African specificity of the trait-associated variation identified here makes annotation with publicly-available regulatory mapping data challenging. To investigate the regulatory function of rs183868412 in immune cells in African populations we used eQTL mapping data from 200 (100 European and 100 African ancestry) individuals in primary monocytes with and without innate stimulation (***Quach et al., 2016***); influenza A virus, LPS, Pam3CSK4 (synthetic lipoprotein and TLR1/2 agonist) and R848 (a TLR7/8 agonist). In this dataset, we found no evidence for a regulatory effect of rs183868412 at the gene level in monocytes regardless of stimulation. We did, however, observe an effect of rs183868412 genotype on expression of a 12bp *BIRC6* exonic sequence (chr2:32,453,943-32,453,954, *p* = 4.40 × 10^−^5), with evidence for co-localisation of this eQTL with our GWAS signal (posterior probability of colocalisation, PP4 = 0.937, Figure 5). This effect was only observed following stimulation with Pam3CSK4 (Figure 5), with the bacteraemia risk allele, rs183868412:T, being associated with reduced expression of this sequence. That 12bp sequence is associated with an alternative splicing event that results in extension of a *BIRC6* exon. The 23rd exon (ENSE00001189810, chr2:32,453,808-32,453,942) of the canonical *BIRC6* transcript, ENST00000421745.6, is 135bp long and terminates immediately before the 12bp sequence associated with rs183868412:T genotype. The 22nd exon (ENSE00003835010, chr2:32,453,808-32,453,942) of an alternative *BIRC6* transcript, ENST00000648282.1, is 147bp long, having the same start site but including the 12bp sequence at its 3’ end. Thus, increased risk of invasive bacterial disease may be associated with decreased expression of an alternative *BIRC6* transcript in TLR1/2 stimulated monocytes.

**Figure 5.**
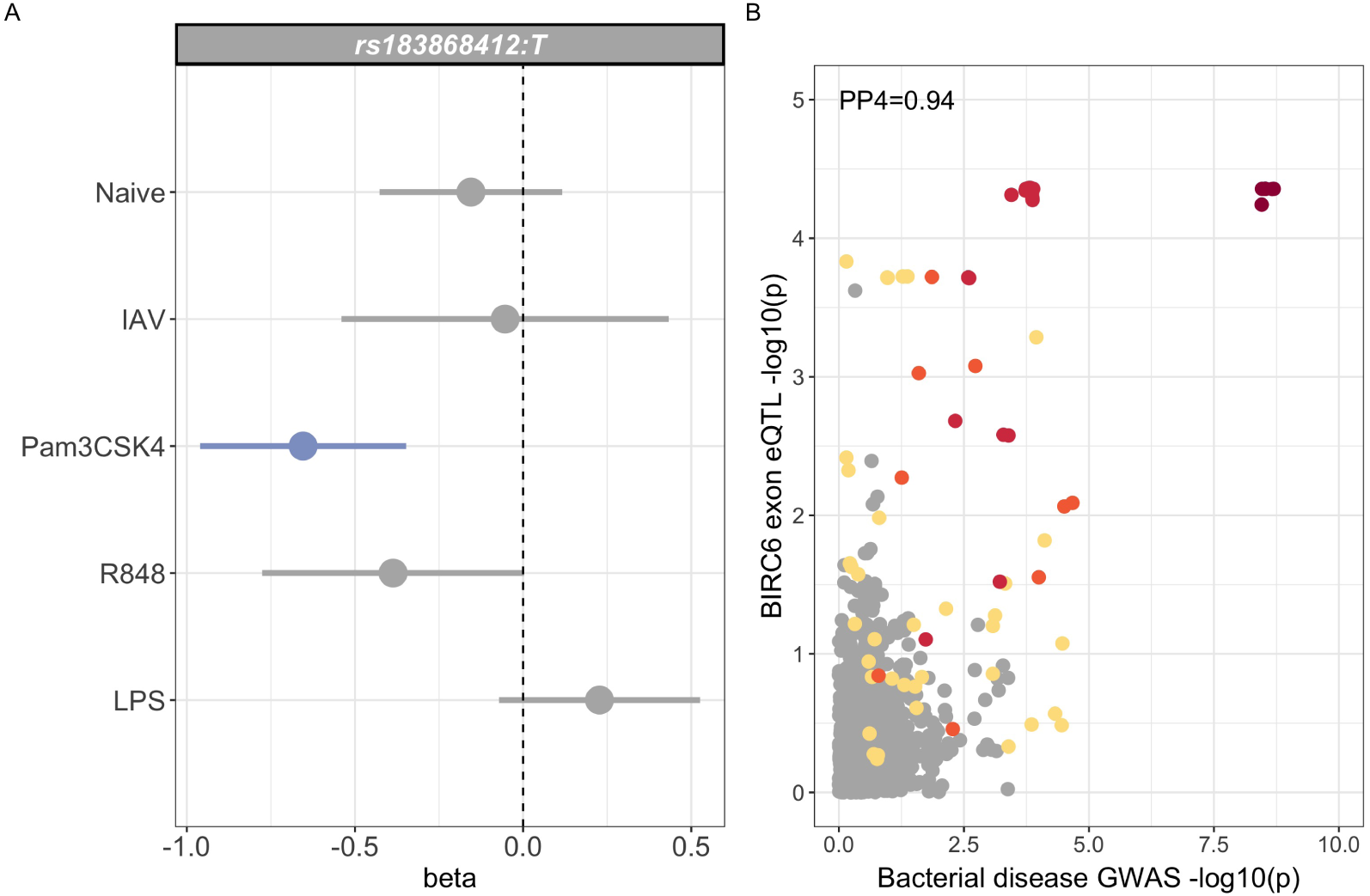
Regulatory function of rs183868412 monocytes. (A) Betas and 95% confidence intervals of rs183868412 association with expression of a 12bp *BIRC6* exonic sequence (chr2:32,453,943-32,453,954) in monocytes. Monocytes are naive or stimulated with; LPS (lipopolysaccharide), IAV (influenza A virus), Pam3CSK4 and R848. (B) Colocalisation of the *BIRC6* exon eQTL in Pam3CSK4-stimulated monocytes colocalizes with the risk locus for invasive bacterial disease (*P P* 4 = 0.937). SNPs are coloured according to linkage disequilibrium to rs183868412.

## Discussion

In this study, we have leveraged the close relationship between *P. falciparum* infection and bacteraemia in African children (***Scott et al., 2011***) to perform a GWAS of invasive bacterial infection in 5,400 Kenyan children. We approached this by defining the probability with which each critically unwell child with a clinical diagnosis of severe malaria has a disease process directly mediated by malaria, that is ‘true’ severe malaria. We hypothesised that critically unwell children, with a low probability of having ‘true’ severe malaria, are enriched for invasive bacterial infections. We explored the validity of this approach, demonstrating that children with a low probability of ‘true’ severe malaria were indeed enriched for culture-proven bacteraemia and were at a higher risk of death than children with a higher probability. We therefore performed a GWAS weighting cases according to their likelihood of invasive bacterial disease. In doing so, we have identified and validated *BIRC6* as a novel genetic susceptibility locus for all-cause invasive bacterial disease in Kenyan children.

The disease-associated locus modifies risk of invasive bacterial disease caused by a broad range pathogens, including *β*-haemolytic *Streptococci, E. coli*, nontyphoidal *Salmonella, S. pneumoniae* and *S. aureus*. Moreover it modifies risk of invasive infection in both the neonatal period and in older children. Furthermore, in contrast to the rs334 *HBB* A>T mutation (***Scott et al., 2011***), rs183868412 modifies risk of invasive bacterial disease in a manner independent of malaria, with rs183868412:T carriage increasing risk of disease across a period of falling malaria transmission and in children with and without concurrent parasitaemia.

We further demonstrate that rs183868412 mediates risk of invasive bacterial disease through the modification of *BIRC6* splicing in Pam3CSK4-stimulated monocytes. *BIRC6* (Baculovirus inhibitor of apoptosis protein repeat containing 6), also known as *BRUCE* (BIR repeat containing ubiquitin-conjugating enzyme), encodes a large member of the inhibitor of apoptosis protein (IAP) family (***Hauser et al., 1998***). Members of the IAP family bind to cognate caspases, inhibiting their activity, and thereby cell death, through occlusion of their active site (***Verhagen et al., 2001***). A proportion of IAPs also contain an E3 ubiquitin ligase allowing both direct inhibition of caspases and targeting of caspases for proteasomal degradation (***Verhagen et al., 2001***). *BIRC6* contains both inhibitor of apoptosis domains and an E2/E3 ubiquitin ligase, which function to inhibit apoptosis in response to a variety of stimuli, both by interaction with and degradation of caspase-9, but also through the ubiquitination and degradation of SMAC, an IAP antagonist (***Hao et al., 2004***; ***Bartke et al., 2004***). *BIRC6* also regulates autophagosome-lysosome fusion (***Ebner et al., 2018***), and ubiquitinates (and targets for degradation) LC3, a key effector of autophagosome formation. Thus, *BIRC6* also acts as a negative regulator of autophagy (***Jia and Bonifacino, 2019***).

It is highly plausible that *BIRC6* could determine susceptibility to invasive infection through either its regulation of apoptosis or autophagy (or both). Sepsis induces marked changes in apoptosis across a range of immune cells (***Hotchkiss and Nicholson, 2006***). There is markedly enhanced apoptosis in both dendritic cells (***Hotchkiss et al., 2002***) and in lymphocytes. Enhanced lymphocyte apoptosis is most striking in B cells and CD4^+^ T cells (***Hotchkiss et al., 2001***) which, at least in part, is mediated by caspase-9. The consequent lymphopoenia is correlated with both severity of sepsis and outcome (***Le Tulzo et al., 2002***). In addition to the direct effects of immune cell loss on the innate and adaptive immune responses to invasive infection, sepsis-induced apoptosis induces immune cell dysfunction, phagocytosis of apoptotic cells resulting in reduction in pro-inflammatory cytokine production and cross-presentation of antigen from apoptotic cells to adaptive immune cells (***Albert, 2004***). In keeping with a role for regulators of apoptosis in the pathogenesis of sepsis, members of the IAP family, including *NAIP/BIRC1* and *BIRC3*, are downregulated in immune cells in patients with sepsis, as is the *BIRC6* ubiquitination target *SMAC* (***Hoogerwerf et al., 2010***). Autophagy contributes to the direct removal of intracellular pathogens and, through the degradation of invading organisms in autophagosomes, directs antigen presentation and pro-inflammatory cytokine secretion (***Deretic et al., 2013***). As above, *BIRC6* regulates autophagy through its interaction with LC3, and overexpression of *LC3B* limits inflammation and tissue injury in a mouse model of sepsis (***Lo et al., 2013***).

In keeping with a role for *BIRC6* in autophagy and apoptosis in sepsis, our data identify a role for genetic variation at *BIRC6* in determining risk of invasive infection secondary to a broad range of bacteria. This is in contrast to previously-published data describing susceptibility to invasive bacterial infection, which has highlighted a prominent role for genetic risk factors that are specific to single pathogens (***Davila et al., 2010***; ***Gilchrist et al., 2018***; ***Rautanen et al., 2016***). In this study, the derived allele (T) at rs183868412 was associated with increased risk of bacteraemia secondary to gram positive (*β*-haemolytic *Streptococci, S. pneumoniae* and *S. aureus*) and gram negative (*E. coli*, nontyphoidal *Salmonella*) pathogens, including intracellular and extracellular bacteria, and enteric and respiratory pathogens. Moreover, rs183868412 modified risk of bacteraemia in both the neonatal period, when infection is likely to be maternally derived, and in older children, when sources of community acquired infection will be more diverse. This modulation of invasive bacterial disease risk, despite diverse sources and routes of infection and diverse mechanisms of invasion, suggests a mechanism in which genetic variation at *BIRC6* modifies risk of invasive infection downstream of initial mechanisms of infection and invasion. The African-specific nature of the trait-associated variation identified here limits our ability to comprehensively interrogate the effect of that variation in immune cells. The eQTL mapping data that we utilise here (***Quach et al., 2016***) is limited in that it allows us only to consider a single immune cell type. A more complete understanding of the role played by genetic variation at *BIRC6* plays in the pathogenesis of sepsis in African children will require more detailed expression and functional studies in African populations.

Taken together, our data identify a role for *BIRC6* in the pathogenesis of invasive bacterial infections in Kenyan children. By maximising our available sample size to include children with a high likelihood of invasive bacterial infection, but without culture-confirmed infection, we facilitate novel variant discovery and reveal a common genetic architecture of invasive bacterial disease secondary to diverse pathogens. In doing so we expand our understanding of the biology of invasive infection in African children. In particular, these data inform our understanding of the biology shared by diverse bacterial infections causing a common clinical syndrome: sepsis.

## Materials and Methods

### Study samples

Recruitment of the severe malaria cases, bacteraemia cases and healthy controls have been described in detail elsewhere (***Ndila et al., 2018***; ***Rautanen et al., 2016***). In brief, children under 14 years admitted to the high dependency ward of Kilifi County Hospital with a clinical diagnosis of severe malaria, defined as *P. falciparum* parasites on blood film and at least one of; reduced Blantyre Coma Score, severe anaemia (Hb<50g/L), evidence of respiratory distress, hypoglycaemia or hyperparasitaemia were eligible for recruitment as cases of severe malaria. During the study period, all children admitted to hospital, with the exception of elective surgical admissions and minor trauma, had a blood sample taken for bacterial culture (BACTEC 9050 instrument, Becton Dickinson, USA). Children under 14 years, in whom a pathogenic organism was identified in blood were eligible for study inclusion (*Bacillus* species, coryneform bacteria, coagulase-negative *Staphyococci, Staphylococcus saprophyticus* and Viridans group *Streptococci* were considered contaminants). Control children were recruited between 3 and 12 months of age from consecutive live births from the population which Kilifi County Hospital serves. Control children have been subject to longitudinal follow-up.

### Models to define the probability of ‘true’ severe malaria

Among cases of severe malaria recruited to the study, we used probabilistic models to assign a probability of that child’s clinical presentation being mediated by parasite sequestration, that is ‘true’ severe malaria, as described previously (***Watson et al., 2021a***,b). Where available (n=1,400), we used platelet counts and *Pf* HRP2 concentrations to derive the probabilistic model. In cases where *Pf* HRP2 concentration was not measured (n=800), we used white blood cell counts and platelet counts as input data to the model.

For both models (Model 1: *Pf* HRP2/platelet counts; Model 2: platelet counts/white blood cell counts), the probabilities of ‘true’ severe malaria were derived by fitting parametric latent class models. These assumed that each patient had a binary latent state (true severe malaria versus not severe malaria). For the Model 1, we assumed that in each latent state the data were distributed as a single bivariate normal distribution. For the model 2, the data did not fit well to a two-component bivariate normal (white blood cell counts have much weaker diagnostic value) so we assumed that the data had bivariate student-t distribution for the severe malaria state, and a flexible mixture of bivariates for the not severe malaria state.

### Genotyping & imputation

For the discovery analysis, we utilised genotypes generated as part of genome-wide association studies of severe malaria (***Band et al., 2019***) and bacteraemia (***Rautanen et al., 2016***) previously reported in this population. Bacteraemia cases and controls were genotyped using the Affymetrix SNP 6.0 array and the severe malaria samples using the Illumina Omni 2.5 M platform. SNP and sample quality control for both datasets are highly analogous, and have been described previously (***Band et al., 2019***; ***Rautanen et al., 2016***). In brief, MalariaGEN SNP QC excluded poorly genotyped SNPs using the following metrics; SNP missingness >2.5%, minor allele frequency (MAF) <1%, Hardy-Weinberg equilibrium (HWE) *p <* 1 × 10^−20^, plate effect *p <* 1 × 10^−3^ and a recall test quantifying changes in genotype following a re-clustering process *p <* 1 × 10^−6^. For Affymetrix-genotyped samples, SNP QC excluded SNPs outside the following QC thresholds; SNP missingness >2%, MAF<1%, genotype probability (info) >0.975, plate effect *p <* 1 × 10^−6^, and HWE *p <* 1 × 10^−20^. Sample QC on both platforms excluded sample outliers with respect to channel intensity, missingness, heterozygosity, population outliers and duplicated samples (IBD>0.75). In addition, for Affymetrix-genotyped samples, samples were excluded in cases of discordant sex as recorded in the clinical record and imputed from mean intensities from X and Y chromosome markers.

To facilitate combining datasets we applied an additional set of cross-platform QC procedures. We defined a shared subset of SNPs genotyped and passing SNP QC on both platforms (n=167,108), observing high levels of genotype concordance (median concordance 0.993, Figure 2–figure supplement 1) among the subset of samples genotyped on both platforms (n=1,365). We used this shared SNP set to compute relatedness estimates and PCs in PLINK v1.90 (***Chang et al., 2015***). The major six PCs of shared genotypes differentiate self-reported ethnicity (Figure 2–figure supplement 2) and are non-differential with respect to genotyping platform (Figure 2–figure supplement 3). To harmonise QC across both platforms we excluded MalariaGEN samples with discordant clinical and genetic sex (n=136). We further excluded one of duplicate or related sample pairs (IBD>0.2) across platforms, retaining case samples where possible and excluding equal numbers of control samples genotyped on each platform (n=1,973). Following QC, genotypes were phased using SHAPEIT2 (***Delaneau et al., 2012***), and untyped genotypes imputed genome-wide using IMPUTE2 (v2.3.2) (***Howie et al., 2011, 2009***) with 1000 Genomes Phase III as a reference panel. Following imputation, we excluded SNPs with imputation info scores <0.5, MAF <1% and HWE *p <* 1 × 10^−5^, applying each threshold for each platform and overall. Following SNP and sample QC, 14,010,600 autosomal SNPs and indels from 5,400 samples (1,445 bacteraemia cases, 1,143 severe malaria cases and 2,812 control samples: 917 Illumina genotyped and 1,895 Affymetrix genotyped) were taken forward as a combined discovery dataset for association analysis.

### Association analysis & fine mapping

In the discovery analysis, we tested for association between genotype at each variant passing QC and invasive bacterial disease by logistic regression in an additive linear model. We used weighted logistic regression to reflect the probability of each case sample being a ‘true’ case of invasive bacterial infection. Cases of culture-confirmed bacteraemia we assigned a weight of 1, whereas cases of severe malaria were assigned weights 1-P(SM|Data), re-weighting the contribution of a case to the log-likelihood according to its probability of representing invasive bacterial infection. Control samples were assigned a weight of 1. Our regression thus assumes that the lower the probability of ‘true’ severe malaria, the greater the probability that a case represents culture-negative invasive bacterial disease. To control for confounding variation, we included the 6 major PCs of genotyping data and genotyping platform as covariates in the model. Weighted logistic regression was implemented using the *glm* function in R. As described previously (***Watson et al., 2021a***), standard errors were transformed to reflect the reduced effective sample size resulting from inclusion of sample weights in the model. We considered *p <* 5 × 10^−8^ to be significant.

We used a Bayesian approach to identify a set of SNPs with 95% probability of containing the causal variant at the trait-associated locus. Approximate Bayes’ factors (***Wakefield, 2009***) were calculated for each SNP in the region (a 200kb surrounding rs183868412) with a prior distribution of *N*(0, 0.2^2^). All SNPs were considered equally likely to be the causal variant a priori. A set of SNPs with 95% probability of containing the causal SNP was defined as the smallest number of SNPs for which the summed posterior probabilities exceed 0.95.

### Replication samples and analysis

To replicate our findings from the discovery analysis we used a second sample set, recruited from the same population as the discovery samples. Replication case samples were cases of bacteraemia only, and did not include cases of severe malaria without culture-confirmed bacterial infection. Case samples were recruited between 1st August 1998 and 30th October 2010. As for the discovery case samples, children under 14 years with a bacterial pathogen isolated from blood on admission to hospital were eligible for recruitment to the study. As above, control samples were recruited as part of a birth cohort from the same population, with children recruited between the ages of 3 and 12 months. Genotyping and QC procedures for these samples have been described previously. In brief, study samples were genotyped using the Immunochip Consortium (***Cortes and Brown, 2011***) array (Illumina). Sample QC was performed as for the discovery samples (above), with duplicate control samples (samples common to MalariaGEN and Immunochip controls, n=78) being removed from the replication set. As above, relatedness estimates and PCs were computed in PLINK v1.90 (***Chang et al., 2015***), with the major four PCs of genotype data differentiating self-reported ethnicity (Figure 3-figure supplement 1). SNP QC excluded the following variants; SNP missingness >1%, MAF <1% and HWE *p <* 1 × 10^−10^. Following QC, 143,000 genotyped variants in 434 cases and 1,258 control samples were taken forward for imputation. As above, imputation was performed with SHAPEIT2 (***Delaneau et al., 2012***) and IMPUTE2 (v2.3.2) (***Howie et al., 2011, 2009***) with 1000 Genomes Phase III as a reference panel.

Following imputation, we further excluded poorly-imputed (imputation info score<0.5) and rare (MAF<1%) variants and variants with HWE *p <* 1×10^−10^. At variants associated with invasive bacterial disease (*p <* 5 × 10^−8^) in the discovery analysis, we tested for association with bacteraemia case-control status using logistic regression in an additive model in SNPTEST v2.5.6 (***Marchini et al., 2007***). To exclude confounding variation, we included the major four PCs of genotyping data in the model. We considered evidence of association with bacteraemia in the replication samples with *p <* 0.05 with the same direction of effect as in the discovery analysis to be significant.

### Bayesian comparison of models of association

At the locus of interest, we used multinomial logistic regression, implemented in SNPTEST v2.5.6 (***Marchini et al., 2007***) to estimate the additive effect of genotype on risk of bacteraemia stratified by pathogen, bacteraemia in the neonatal and non-neonatal periods, bacteraemia with and without *P. falciparum* parasitaemia, and bacteraemia presenting at different time periods across a period of declining malaria transmission intensity. For these analyses we used only samples with culture-confirmed bacteraemia. In each case we used control status as the baseline stratum, and included the major principal components of genotyping data to control for confounding variation as above.

For the pathogen-stratified analysis, we defined eight case strata among the discovery cases, one for each of the seven most commonly isolated organisms (*Acinetobacter*, n=118; *β*-haemolytic *Streptococci*, n=130; *E. coli*, n=141; *H. in!uenzae* type b, n=113; nontyphoidal *Salmonella*, n=159; pneumococci, n=390; *S. aureus*, n=152) and one stratum for the remaining other organisms (n=242). For the neonatal/non-neonatal disease analysis we stratified cases as presenting in the first 28 days of life (n=195) or beyond that (n=1,245). For the analysis comparing bacteraemia with and without malaria we stratified cases with (n=204) and without (n=1,236) *P. falciparum* on their admission blood film. For each of these analysis case strata were compared to Affymetrix-genotyped discovery control samples (n=1,895) as a baseline stratum.

For the analysis stratified across year of admission, we defined case strata by grouping into six time periods according to their date of admission; 1998-2000 (n=498), 2001-2002 (n=349), 2003-2004 (n=467), 2005-2006 (n=310), 2007-2008 (n=237), 2009-2010 (n=111). For this analysis we used both discovery (Affymetrix) and replication (Immunochip) case and control samples. This allowed better coverage of the years later in the study, which were underrepresented in the discovery samples (the discovery median admission year is 2003, c.f. 2005 for the replication samples). In that analysis we used multinomial logistic regression in each cohort to estimate stratum-specific effects, combining these results in a fixed effects meta-analysis using BINGWA (***Band et al., 2015***). We then compared models of association using a Bayesian approach (***Rautanen et al., 2016***), considering the following models:

“Null”: effect size = i.e. no association with bacteraemia.

“Same”: effect size *N*(0, 0.2^2^) and fixed across strata.

Additional models consider each possible combination of a fixed effect size for associated strata and no association at other strata. For each model we calculated approximate Bayes factors (***Wakeield, 2009***) and posterior probabilities, assuming each model to be equally likely a priori. Statistical analysis was performed in R.

### Co-localisation analysis

We used the R package coloc v5.1.0 (***Giambartolomei et al., 2014***) to identify evidence of causal variants shared by our bacterial disease-associated locus of interest and regulatory genetic variation mapped by eQTL studies. We downloaded eQTL summary statistics describing eQTL mapping in naive and stimulated monocytes (***Quach et al., 2016***), including study participants with African ancestry, from the eQTL catalogue (***Kerimov et al., 2021***) (https://github.com/eQTL-Catalogue/eQTL-Catalogue-resources/, accessed 28th October 2021). Coloc adopts a Bayesian approach to compare evidence for independent or shared association signals for two traits at a given genetic locus. We used the coloc.susie() command to allow co-localisation of multiple independent signals at a single locus for each trait. We tested for colocalisation between invasive bacterial disease susceptibility at the *BIRC6* locus and previously-published eQTL data in naive and stimulated primary monocytes isolated from individuals of European (n=100) and African (n=100) ancestry (***Quach et al., 2016***). We considered evidence for co-localisation for each gene and exon within a 250kb window of the peak association (rs183868412). We considered a posterior probability > 0.8 supporting a shared causal locus to be significant.

## Data Availability

Patient level genotype and phenotype data are available via the European Genome-Phenome Archive, with accession codes EGAD00010000950 (WTCCC2: bacteraemia cases and controls) and EGAD00010000904 (MalariaGEN Consortium: severe malaria cases and controls). Full GWAS summary statistics have been deposited with the GWAS Catalog with accession code GCST90094632.

## Acknowledgments

This publication uses genotyping data from the MalariaGEN consortial project, as described in Malaria Genomic Epidemiology Network, et al. Nature Communications, 2019 (https://doi.org/10.1038/s41467-019-13480-z). This study makes use of data generated by the Wellcome Trust Case Control Consortium 2 project (Grant Reference 085475/B/08/Z). JJG and AJM are funded by National Institute for Health Research (NIHR) Clinical Lectureships. TNW and JAGS are supported by Senior Research Fellowships from the Wellcome Trust (202800 and 098532 respectively). JAW is a Sir Henry Dale Fellow funded by the Wellcome Trust (223253/Z/21/Z). SMAART (Severe Malaria Africa – A consortium for Research and Trials) is funded by a Wellcome Collaborative Award in Science grant (209265/Z/17/Z) held in part by KM and TNW. During this work AVSH was supported by a Wellcome Trust Senior Investigator Award (HCUZZ0) and by a European Research Council advanced grant (294557). The research was supported by the Wellcome Trust Core Award Grant Number 203141/Z/16/Z with additional support from the NIHR Oxford BRC. The views expressed are those of the author(s) and not necessarily those of the NHS, the NIHR or the Department of Health. This research was funded by The Wellcome Trust. A CC BY or equivalent licence is applied to the author accepted manuscript arising from this submission, in accordance with the grant’s open access conditions. This paper is published with the permission of the Director of KEMRI.

---

**Figure 1–Figure supplement 1.**
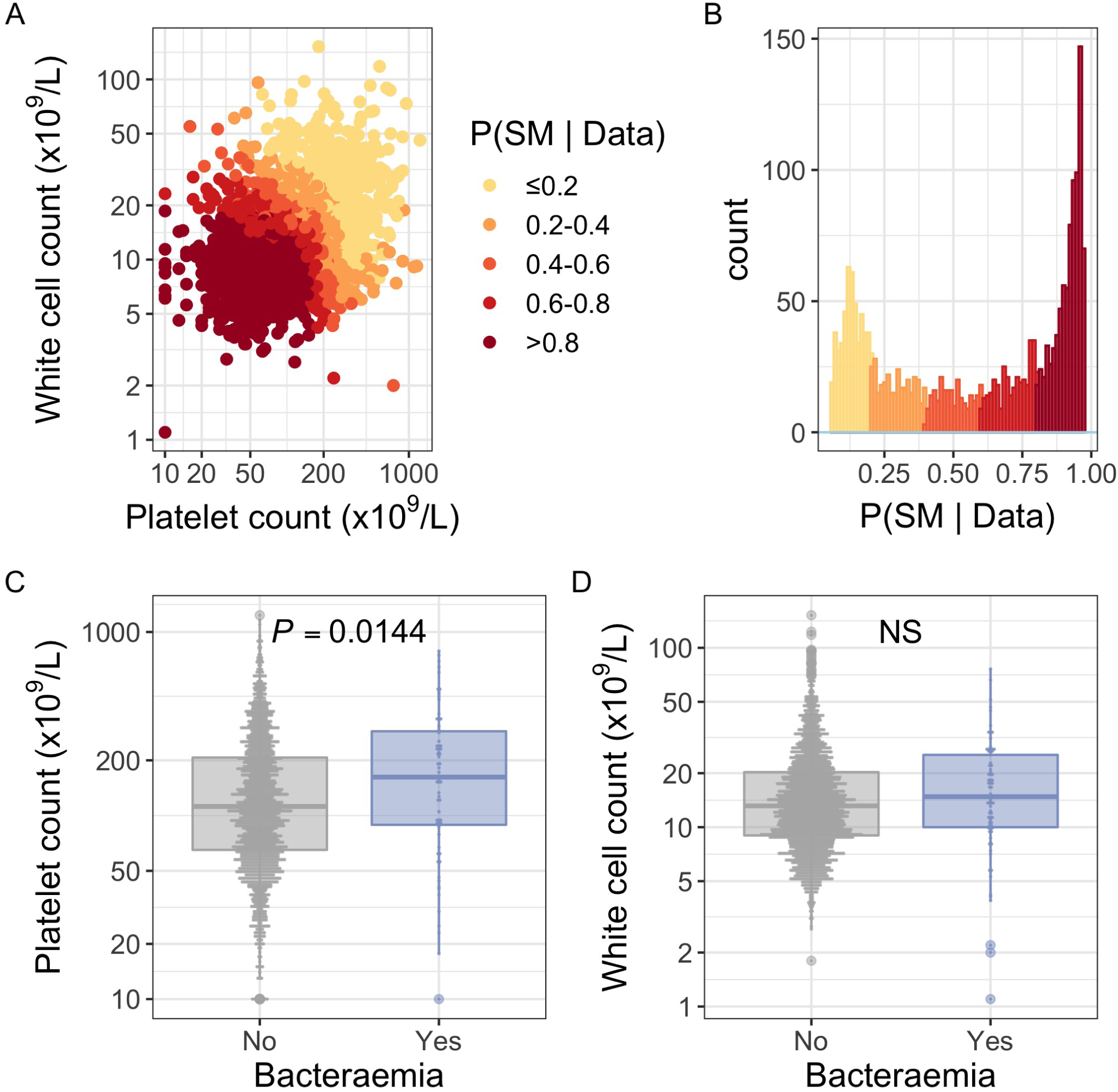
White blood cell and platelet count as predictors of severe malaria. (A) Distribution of white blood cell and platelet count among Kenyan children (n=2,200) with a clinical diagnosis of severe malaria. Points are coloured according to the probability of ‘true’ severe malaria given the data. (B) Distribution of ‘true’ severe malaria probabilities estimated from platelet count and white blood cell count. (C) Platelets counts in children with a clinical diagnosis of severe malaria with and without concomitant bacteraemia. (D) White blood cell counts in children with a clinical diagnosis of severe malaria with and without concomitant bacteraemia.

**Figure 2–Figure supplement 1.**
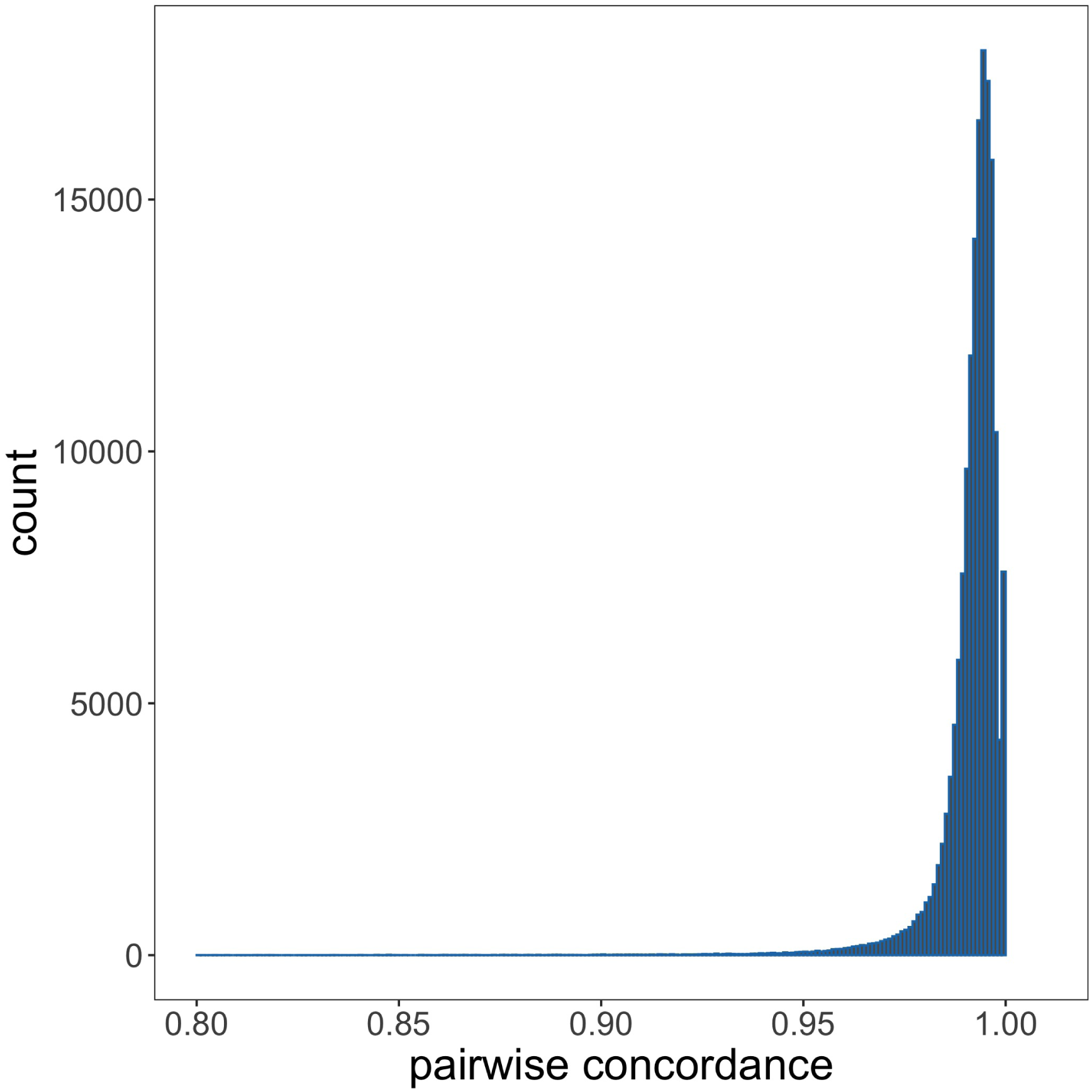
Genotyping concordance between Illumina and Affymetrix platforms. Pairwise genotyping concordance between samples genotyped on both Affymetrix SNP 6.0 and Illumina Omni 2.5M platforms.

**Figure 2–Figure supplement 2.**
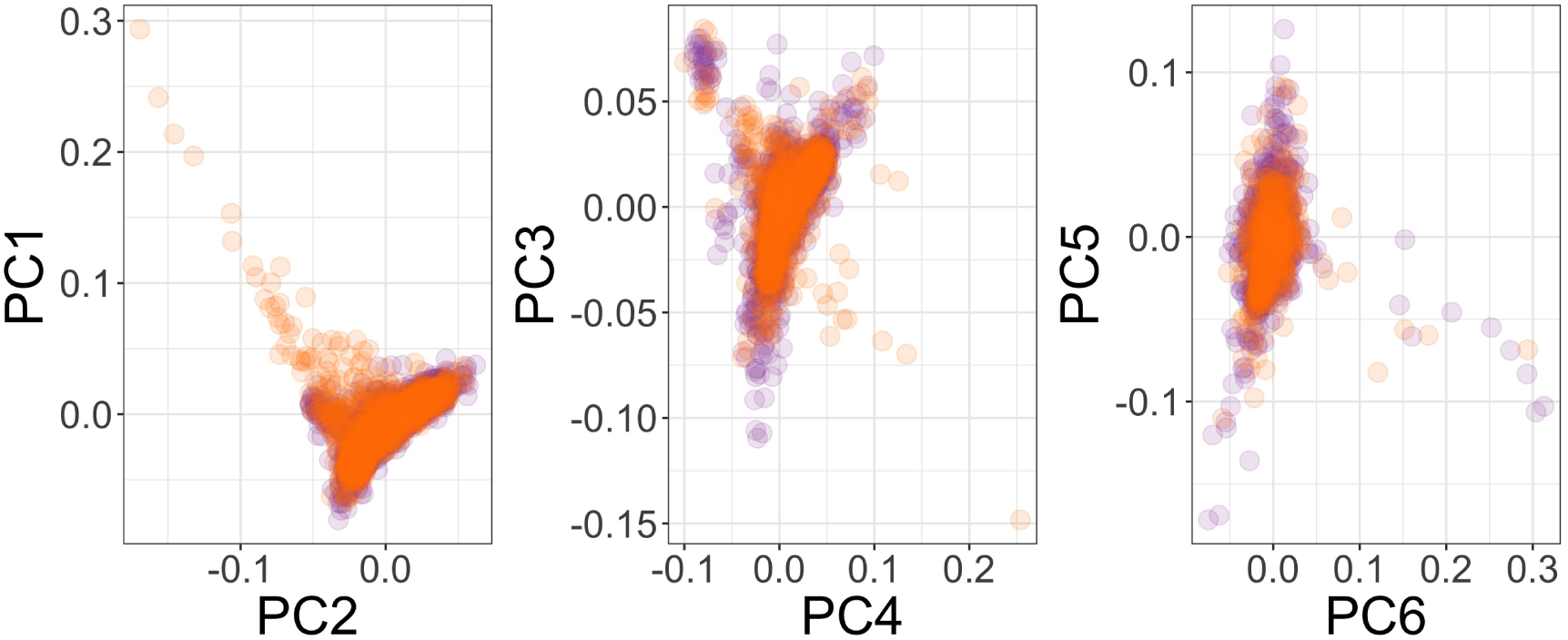
Principal components of genome-wide genotyping data in discovery samples. Individuals are color-coded according to genotyping platform; Affymetrix SNP 6.0 in purple, Illumina Omni 2.5M in orange.

**Figure 2–Figure supplement 3.**
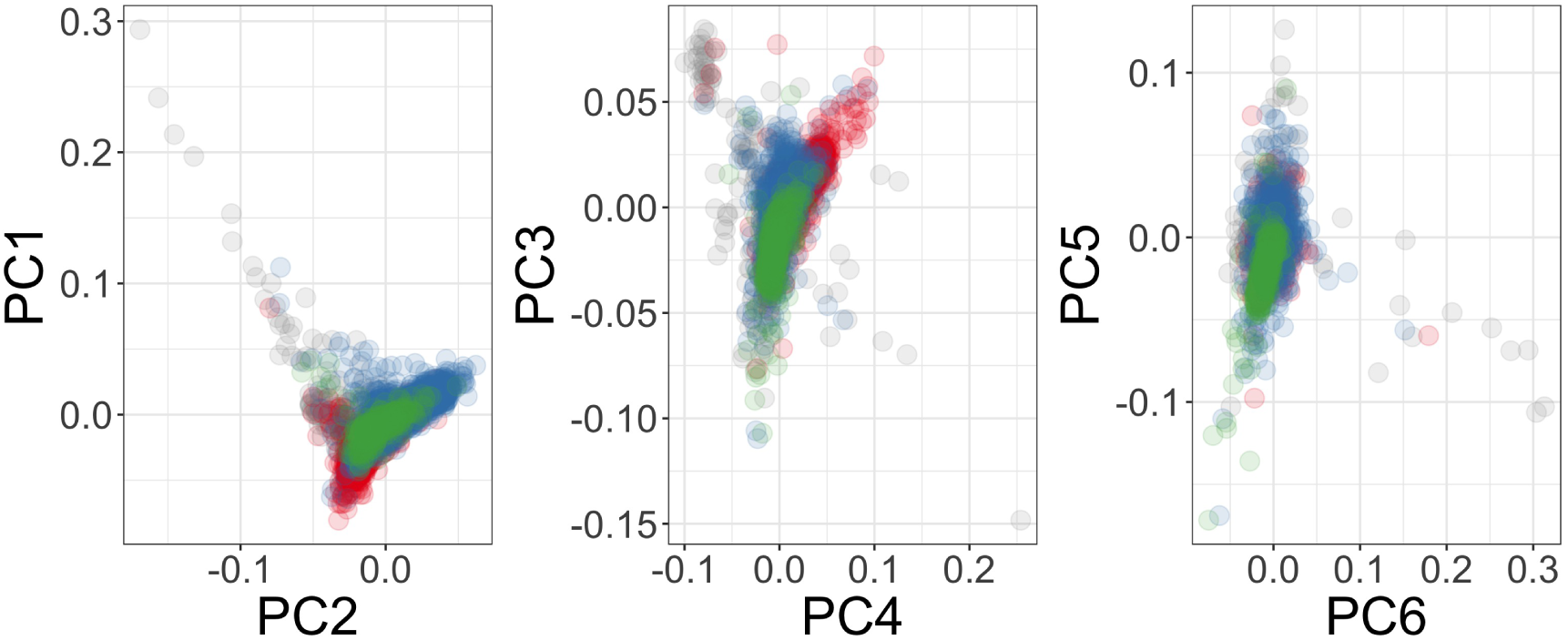
Principal components of genome-wide genotyping data in discovery samples. Individuals are color-coded according to self-reported ethnicity; Chonyi in red, Giriama in blue, Kauma in green and other in grey.

**Figure 2–Figure supplement 4.**
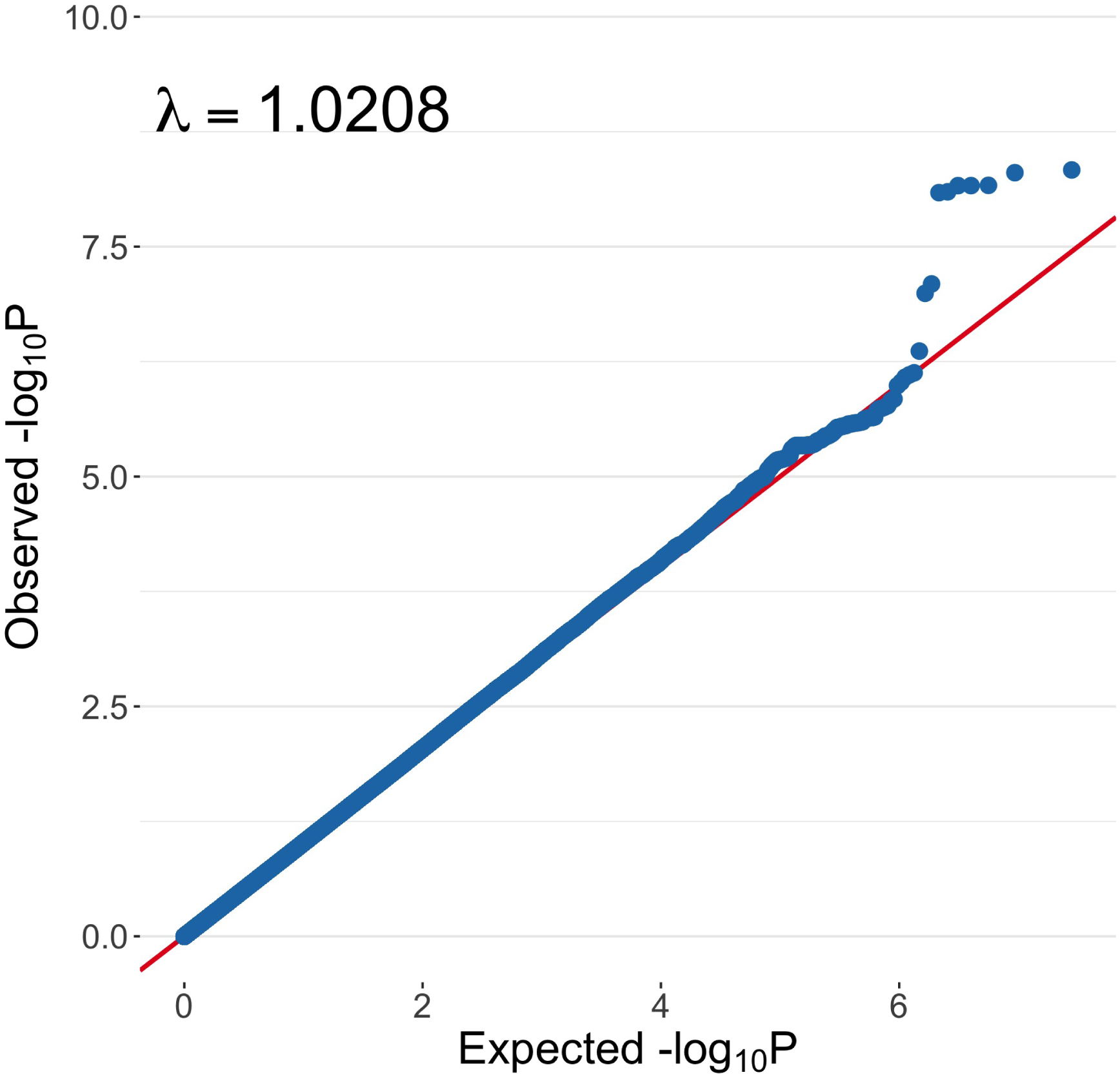
Quantile-quantile plot of invasive bacterial infection in Kenyan children. QQ plot of weighted logistic regression GWAS of invasive bacterial disease in Kenyan children (2,588 cases, 2,812 controls).

**Figure 3–Figure supplement 1.**
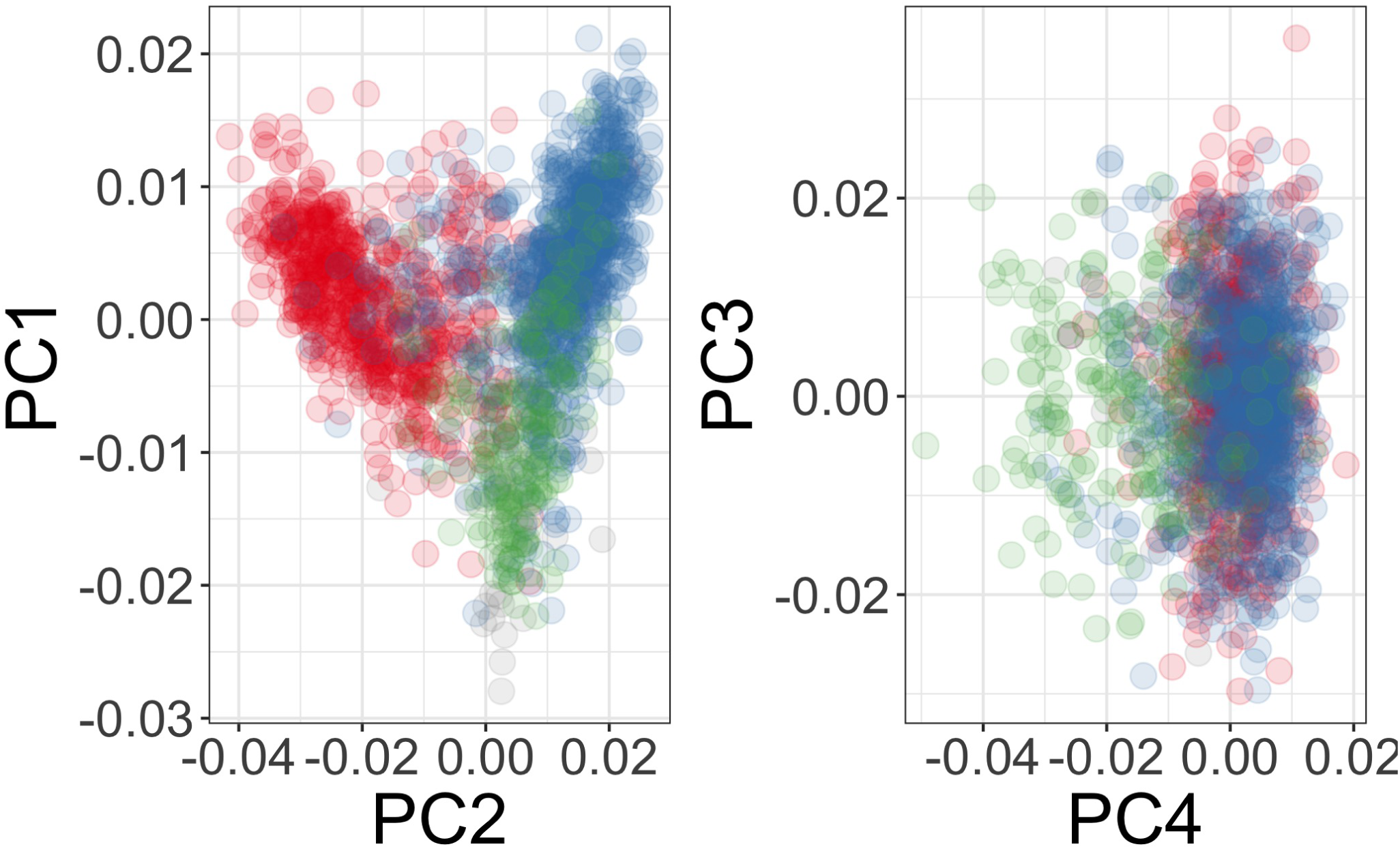
Principal components of genome-wide genotyping data in replication samples. Individuals are color-coded according to self-reported ethnicity; Chonyi in red, Giriama in blue, Kauma in green and other in grey.

## References

Albert ML. Death-defying immunity: do apoptotic cells influence antigen processing and presentation? Nature Reviews Immunology. 2004; 4(3):223–231. https://doi.org/10.1038/nri11308, doi: 10.1038/nri11308.

Band G, Le QS, Clarke GM, Kivinen K, Hubbart C, Jeffreys AE, Rowlands K, Leffler EM, Jallow M, Conway DJ, Sisay-Joof F, Sirugo G, d’Alessandro U, Toure OB, Thera MA, Konate S, Sissoko S, Mangano VD, Bougouma EC, Sirima SB, et al. Insights into malaria susceptibility using genome-wide data on 17,000 individuals from Africa, Asia and Oceania. Nature Communications. 2019; 10(1):5732. https://doi.org/10.1038/s41467-019-13480-z, doi: 10.1038/s41467-019-13480-z.

Band G, Rockett KA, Spencer CCA, Kwiatkowski DP, Si Le Q, Clarke GM, Kivinen K, Leffler EM, Rockett KA, Kwiatkowski DP, Spencer CCA, Rockett KA, Spencer CCA, Cornelius V, Conway DJ, Williams TN, Taylor T, Kwiatkowski DP, Conway DJ, Bojang KA, et al. A novel locus of resistance to severe malaria in a region of ancient balancing selection. Nature. 2015; 526(7572):253–257. https://doi.org/10.1038/nature15390, doi: 10.1038/nature15390.

Bartke T, Pohl C, Pyrowolakis G, Jentsch S. Dual Role of BRUCE as an Antiapoptotic IAP and a Chimeric E2/E3 Ubiquitin Ligase. Molecular Cell. 2004; 14(6):801–811. https://www.sciencedirect.com/science/article/pii/S1097276504003041, doi: https://doi.org/10.1016/j.molcel.2004.05.018.

Bejon P, Berkley JA, Mwangi T, Ogada E, Mwangi I, Maitland K, Williams T, Scott JAG, English M, Lowe BS, Peshu N, Newton Crjc, Marsh K. Defining childhood severe falciparum malaria for intervention studies. PLoS Med. 2007 Aug; 4(8):e251. doi: 10.1371/journal.pmed.0040251.

Berkley JA, Lowe BS, Mwangi I, Williams T, Bauni E, Mwarumba S, Ngetsa C, Slack MPE, Njenga S, Hart CA, Maitland K, English M, Marsh K, Scott JAG. Bacteremia among children admitted to a rural hospital in Kenya. N Engl J Med. 2005 Jan; 352(1):39–47. doi: 10.1056/NEJMoa040275.

Bustamante J, Boisson-Dupuis S, Abel L, Casanova JL. Mendelian susceptibility to mycobacterial disease: genetic, immunological, and clinical features of inborn errors of IFN- immunity. Semin Immunol. 2014 Dec; 26(6):454–470. doi: 10.1016/j.smim.2014.09.008.

Chang CC, Chow CC, Tellier LC, Vattikuti S, Purcell SM, Lee JJ. Second-generation PLINK: rising to the challenge of larger and richer datasets. Gigascience. 2015; 4:7. doi: 10.1186/s13742-015-0047-8.

Church J, Maitland K. Invasive bacterial co-infection in African children with Plasmodium falciparum malaria: a systematic review. BMC Med. 2014 Feb; 12:31. doi: 10.1186/1741-7015-12-31.

Cortes A, Brown MA. Promise and pitfalls of the Immunochip. Arthritis Res Ther. 2011 Feb; 13(1):101. doi: 10.1186/ar3204.

Cowgill KD, Ndiritu M, Nyiro J, Slack MPE, Chiphatsi S, Ismail A, Kamau T, Mwangi I, English M, Newton Crjc, Feikin DR, Scott JAG. Effectiveness of Haemophilus influenzae Type b Conjugate Vaccine Introduction Into Routine Childhood Immunization in Kenya. JAMA. 2006 11/3/2021; 296(6):671–678. https://doi.org/10.1001/jama.296.6.671, doi: 10.1001/jama.296.6.671.

Davila S, Wright VJ, Khor CC, Sim KS, Binder A, Breunis WB, Inwald D, Nadel S, Betts H, Carrol ED, de Groot R, Hermans PWM, Hazelzet J, Emonts M, Lim CC, Kuijpers TW, Martinon-Torres F, Salas A, Zenz W, Levin M, et al. Genome-wide association study identifies variants in the CFH region associated with host susceptibility to meningococcal disease. Nature Genetics. 2010; 42(9):772–776. https://doi.org/10.1038/ng.640, doi: 10.1038/ng.640.

Delaneau O, Marchini J, Zagury JF. A linear complexity phasing method for thousands of genomes. Nature Methods. 2012; 9(2):179–181. https://doi.org/10.1038/nmeth.1785, doi: 10.1038/nmeth.1785.

Deretic V, Saitoh T, Akira S. Autophagy in infection, inflammation and immunity. Nature Reviews Immunology. 2013; 13(10):722–737. https://doi.org/10.1038/nri3532, doi: 10.1038/nri3532.

Driscoll AJ, Deloria Knoll M, Hammitt LL, Baggett HC, Brooks WA, Feikin DR, Kotloff KL, Levine OS, Madhi SA, O’Brien KL, Scott JAG, Thea DM, Howie SRC, Adrian PV, Ahmed D, DeLuca AN, Ebruke BE, Gitahi C, Higdon MM, Kaewpan A, et al. The Effect of Antibiotic Exposure and Specimen Volume on the Detection of Bacterial Pathogens in Children With Pneumonia. Clinical Infectious Diseases. 2017 11/16/2021; 64(Suppl_3):S368– S377. https://doi.org/10.1093/cid/cix101, doi: 10.1093/cid/cix101.

Ebner P, Poetsch I, Deszcz L, Hoffmann T, Zuber J, Ikeda F. The IAP family member BRUCE regulates autophagosome-lysosome fusion. Nat Commun. 2018 Feb; 9(1):599. doi: 10.1038/s41467-018-02823-x.

Figueroa J, Andreoni J, Densen P. Complement deficiency states and meningococcal disease. Immunol Res. 1993; 12(3):295–311. doi: 10.1007/BF02918259.

Giambartolomei C, Vukcevic D, Schadt EE, Franke L, Hingorani AD, Wallace C, Plagnol V. Bayesian test for colocalisation between pairs of genetic association studies using summary statistics. PLoS Genet. 2014 May; 10(5):e1004383. doi: 10.1371/journal.pgen.1004383.

Gilchrist JJ, Rautanen A, Fairfax BP, Mills TC, Naranbhai V, Trochet H, Pirinen M, Muthumbi E, Mwarumba S, Njuguna P, Mturi N, Msefula CL, Gondwe EN, MacLennan JM, Chapman SJ, Molyneux ME, Knight JC, Spencer CCA, Williams TN, MacLennan CA, et al. Risk of nontyphoidal Salmonella bacteraemia in African children is modified by STAT4. Nature Communications. 2018; 9(1):1014. https://doi.org/10.1038/s41467-017-02398-z, doi: 10.1038/s41467-017-02398-z.

Hao Y, Sekine K, Kawabata A, Nakamura H, Ishioka T, Ohata H, Katayama R, Hashimoto C, Zhang X, Noda T, Tsuruo T, Naito M. Apollon ubiquitinates SMAC and caspase-9, and has an essential cytoprotection function. Nature Cell Biology. 2004; 6(9):849–860. https://doi.org/10.1038/ncb1159, doi: 10.1038/ncb1159.

Hauser HP, Bardroff M, Pyrowolakis G, Jentsch S. A giant ubiquitin-conjugating enzyme related to IAP apoptosis inhibitors. J Cell Biol. 1998 Jun; 141(6):1415–1422. doi: 10.1083/jcb.141.6.1415.

Hendriksen ICE, Mwanga-Amumpaire J, von Seidlein L, Mtove G, White LJ, Olaosebikan R, Lee SJ, Tshefu AK, Woodrow C, Amos B, Karema C, Saiwaew S, Maitland K, Gomes E, Pan-Ngum W, Gesase S, Silamut K, Reyburn H, Joseph S, Chotivanich K, et al. Diagnosing severe falciparum malaria in parasitaemic African children: a prospective evaluation of plasma PfHRP2 measurement. PLoS Med. 2012; 9(8):e1001297. doi: 10.1371/journal.pmed.1001297.

Hoogerwerf JJ, van Zoelen MA, Wiersinga WJ, van ‘t Veer C, de Vos AF, de Boer A, Schultz MJ, Hooibrink B, de Jonge E, van der Poll T. Gene expression profiling of apoptosis regulators in patients with sepsis. J Innate Immun. 2010; 2(5):461–468. doi: 10.1159/000317035.

Hotchkiss RS, Tinsley KW, Swanson PE, Schmieg REJ, Hui JJ, Chang KC, Osborne DF, Freeman BD, Cobb JP, Buchman TG, Karl IE. Sepsis-induced apoptosis causes progressive profound depletion of B and CD4+ T lymphocytes in humans. J Immunol. 2001 Jun; 166(11):6952–6963. doi: 10.4049/jimmunol.166.11.6952.

Hotchkiss RS, Nicholson DW. Apoptosis and caspases regulate death and inflammation in sepsis. Nat Rev Immunol. 2006 Nov; 6(11):813–822. doi: 10.1038/nri1943.

Hotchkiss RS, Tinsley KW, Swanson PE, Grayson MH, Osborne DF, Wagner TH, Cobb JP, Coopersmith C, Karl IE. Depletion of dendritic cells, but not macrophages, in patients with sepsis. J Immunol. 2002 Mar; 168(5):2493– 2500. doi: 10.4049/jimmunol.168.5.2493.

Howie B, Marchini J, Stephens M. Genotype imputation with thousands of genomes. G3 (Bethesda). 2011 Nov; 1(6):457–470. doi: 10.1534/g3.111.001198.

Howie BN, Donnelly P, Marchini J. A flexible and accurate genotype imputation method for the next generation of genome-wide association studies. PLoS Genet. 2009 Jun; 5(6):e1000529. doi: 10.1371/journal.pgen.1000529.

Jia R, Bonifacino JS. Negative regulation of autophagy by UBA6-BIRC6-mediated ubiquitination of LC3. Elife. 2019 Nov; 8. doi: 10.7554/eLife.50034.

Kerimov N, Hayhurst JD, Peikova K, Manning JR, Walter P, Kolberg L, Samoviča M, Sakthivel MP, Kuzmin I, Trevanion SJ, Burdett T, Jupp S, Parkinson H, Papatheodorou I, Yates AD, Zerbino DR, Alasoo K. A compendium of uniformly processed human gene expression and splicing quantitative trait loci. Nature Genetics. 2021; 53(9):1290–1299. https://doi.org/10.1038/s41588-021-00924-w, doi: 10.1038/s41588-021-00924-w.

Le Tulzo Y, Pangault C, Gacouin A, Guilloux V, Tribut O, Amiot L, Tattevin P, Thomas R, Fauchet R, Drénou B. Early circulating lymphocyte apoptosis in human septic shock is associated with poor outcome. Shock. 2002 Dec; 18(6):487–494. doi: 10.1097/00024382-200212000-00001.

Lo S, Yuan SSF, Hsu C, Cheng YJ, Chang YF, Hsueh HW, Lee PH, Hsieh YC. Lc3 Over-Expression Improves Survival and Attenuates Lung Injury Through Increasing Autophagosomal Clearance in Septic Mice. Annals of Surgery. 2013; 257(2). https://journals.lww.com/annalsofsurgery/Fulltext/2013/02000/Lc3_Over_Expression_Improves_Survival_and.26.aspx.

Marchini J, Howie B, Myers S, McVean G, Donnelly P. A new multipoint method for genome-wide association studies by imputation of genotypes. Nature Genetics. 2007; 39(7):906–913. https://doi.org/10.1038/ng2088, doi: 10.1038/ng2088.

Ndila CM, Uyoga S, Macharia AW, Nyutu G, Peshu N, Ojal J, Shebe M, Awuondo KO, Mturi N, Tsofa B, Sepúlveda N, Clark TG, Band G, Clarke G, Rowlands K, Hubbart C, Jeffreys A, Kariuki S, Marsh K, Mackinnon M, et al. Human candidate gene polymorphisms and risk of severe malaria in children in Kilifi, Kenya: a case-control association study. Lancet Haematol. 2018 Aug; 5(8):e333–e345. doi: 10.1016/S2352-3026(18)30107-8.

Picard C, von Bernuth H, Ku CL, Yang K, Puel A, Casanova JL. Inherited human IRAK-4 deficiency: an update. Immunol Res. 2007; 38(1-3):347–352. doi: 10.1007/s12026-007-0006-2.

Quach H, Rotival M, Pothlichet J, Loh YHE, Dannemann M, Zidane N, Laval G, Patin E, Harmant C, Lopez M, Deschamps M, Naffakh N, Duffy D, Coen A, Leroux-Roels G, Clément F, Boland A, Deleuze JF, Kelso J, Albert ML, et al. Genetic Adaptation and Neandertal Admixture Shaped the Immune System of Human Populations. Cell. 2016 Oct; 167(3):643–656. doi: 10.1016/j.cell.2016.09.024.

Rautanen A, Pirinen M, Mills TC, Rockett KA, Strange A, Ndungu AW, Naranbhai V, Gilchrist JJ, Bellenguez C, Freeman C, Band G, Bumpstead SJ, Edkins S, Giannoulatou E, Gray E, Dronov S, Hunt SE, Langford C, Pearson RD, Su Z, et al. Polymorphism in a lincRNA Associates with a Doubled Risk of Pneumococcal Bacteremia in Kenyan Children. Am J Hum Genet. 2016 Jun; 98(6):1092–1100. doi: 10.1016/j.ajhg.2016.03.025.

Scott JAG, Berkley JA, Mwangi I, Ochola L, Uyoga S, Macharia A, Ndila C, Lowe BS, Mwarumba S, Bauni E, Marsh K, Williams TN. Relation between falciparum malaria and bacteraemia in Kenyan children: a population-based, case-control study and a longitudinal study. Lancet. 2011 Oct; 378(9799):1316–1323. doi: 10.1016/S01406736(11)60888-X.

Silaba M, Ooko M, Bottomley C, Sande J, Benamore R, Park K, Ignas J, Maitland K, Mturi N, Makumi A, Otiende M, Kagwanja S, Safari S, Ochola V, Bwanaali T, Bauni E, Gleeson F, Deloria Knoll M, Adetifa I, Marsh K, et al. Effect of 10-valent pneumococcal conjugate vaccine on the incidence of radiologically-confirmed pneumonia and clinically-defined pneumonia in Kenyan children: an interrupted time-series analysis. The Lancet Global Health. 2019 2021/11/03; 7(3):e337–e346. https://doi.org/10.1016/S2214-109X(18)30491-1, doi: 10.1016/S2214-109X(18)30491-1.

Verhagen AM, Coulson EJ, Vaux DL. Inhibitor of apoptosis proteins and their relatives: IAPs and other BIRPs. Genome Biology. 2001; 2(7):reviews3009.1. https://doi.org/10.1186/gb-2001-2-7-reviews3009, doi: 10.1186/gb-2001-2-7-reviews3009.

Vos T, Lim SS, Abbafati C, Abbas KM, Abbasi M, Abbasifard M, Abbasi-Kangevari M, Abbastabar H, Abd-Allah F, Abdelalim A, Abdollahi M, Abdollahpour I, Abolhassani H, Aboyans V, Abrams EM, Abreu LG, Abrigo MRM, Abu-Raddad LJ, Abushouk AI, Acebedo A, et al. Global burden of 369 diseases and injuries in 204 countries and territories, 1990–2019: a systematic analysis for the Global Burden of Disease Study 2019. The Lancet. 2020 2021/11/03; 396(10258):1204–1222. https://doi.org/10.1016/S0140-6736(20)30925-9, doi: 10.1016/S0140-6736(20)30925-9.

Wakefield J. Bayes factors for genome-wide association studies: comparison with P-values. Genet Epidemiol. 2009 Jan; 33(1):79–86. doi: 10.1002/gepi.20359.

Watson JA, Ndila CM, Uyoga S, Macharia A, Nyutu G, Mohammed S, Ngetsa C, Mturi N, Peshu N, Tsofa B, Rockett K, Leopold S, Kingston H, George EC, Maitland K, Day NP, Dondorp AM, Bejon P, Williams T, Holmes CC, et al. Improving statistical power in severe malaria genetic association studies by augmenting phenotypic precision. Elife. 2021 Jul; 10. doi: 10.7554/eLife.69698.

Watson JA, Uyoga S, Wanjiku P, Makale J, Nyutu GM, Mturi N, George EC, Woodrow CJ, Day NP, Bejon P, Opoka RO, Dondorp AM, John CC, Maitland K, Williams TN, White NJ. Improving the diagnosis of severe malaria in African children using platelet counts and plasma Pf HRP2 concentrations. medRxiv. 2021; https://www.medrxiv.org/content/early/2021/10/30/2021.10.27.21265557, doi: 10.1101/2021.10.27.21265557.

Williams TN, Uyoga S, Macharia A, Ndila C, McAuley CF, Opi DH, Mwarumba S, Makani J, Komba A, Ndiritu MN, Sharif SK, Marsh K, Berkley JA, Scott JAG. Bacteraemia in Kenyan children with sickle-cell anaemia: a retrospective cohort and case-control study. Lancet. 2009 Oct; 374(9698):1364–1370. doi: 10.1016/S0140-6736(09)61374-X.

